# Cost-effectiveness of targeted geographic versus ring vaccination campaigns in response to an outbreak of Ebola Virus Disease caused by *Orthoebolavirus zairense*

**DOI:** 10.1101/2025.05.22.25328133

**Authors:** Gemma Nedjati-Gilani, Victoria M. Cox, Carl A. B. Pearson, Janetta E. Skarp, Rob Johnson, Dav M. Ebengo, Placide Mbala-Kingebeni, David Niyukuri, Christian Morgenstern, Wes Hinsley, Steven Riley, Abigail Hunter, Ruth McCabe, Aminata Bagayoko, Alejandro Costa, Philipp Lambach, Ruth Kallay, Emma Aberle-Grasse, Sarah Pallas, Pierre Muhoza, Neil M. Ferguson, Lilith K. Whittles, Katharina D. Hauck, Anne Cori

## Abstract

**Background:** WHO recommends ring vaccination (RV) of Ebola virus disease (EVD) case contact chains to contain outbreaks of *Orthoebolavirus zairense*. However, this strategy can falter in remote or conflict-affected settings where effective contact tracing is challenging. Targeted geographic vaccination (TGV) – immunising everyone near an EVD case’s location without requiring contact tracing – has been implemented as an alternative. We compare the epidemiological impact and cost-effectiveness of TGV versus RV under diverse outbreak scenarios.

**Methods:** We extended a spatially-explicit individual-based stochastic EVD transmission model calibrated to outbreaks in the Democratic Republic of the Congo. We simulated outbreaks varying both epidemiological and operational parameters. We evaluated vaccination strategies on total cases, deaths, epidemic duration, vaccine doses used, and incremental net monetary benefit (INMB).

**Results:** No single strategy dominates in all scenarios. For better overall response performance, RV results in substantially fewer cases and deaths than TGV (approximately 20% fewer cases), requires fewer doses, and has higher INMB more frequently. Under pessimistic response assumptions, TGV slightly outperforms RV by preventing more infections, curbing the largest outbreaks, and using fewer doses, although the INMBs are similar between strategies.

**Interpretation:** Different operational contexts recommend different vaccination strategies. Overall, surveillance, contact tracing, and isolation have the most impact on *Orthoebolavirus zairense* outbreak control. Depending on response effectiveness, the preferred strategy for adding vaccination varies. Backed by effective case detection and management, RV appears the better approach. Under severe response constraints, TGV can be more effective.

## 1 Introduction

Since the Ebola Virus Zaire species (*Orthoebolavirus zairense*) was identified in 1976^1^, efforts to develop a vaccine struggled for decades^2^ until the 2014-2016 West African epidemic – which had over 22,000 cases and 11,000 deaths^3^ – when a cluster-randomised ring vaccination (RV) trial in Guinea found a 100% vaccine efficacy against Ebola virus disease (EVD) for rVSV-ZEBOV^4^. Subsequent larger scale evaluation in a more operational setting confirmed high efficacy, estimating around 84%, near the middle of the originally estimated range^5^.

The World Health Organization (WHO) recommended RV – immunising contacts and contacts-of-contacts of confirmed cases – as the primary response strategy^6^, which was deployed under emergency protocols in the 2018-2020 epidemic in Nord Kivu and Ituri in the Democratic Republic of Congo (DRC)^4,7^. In practice, many challenges to contact tracing and ring formation impeded RV: conflict, population mobility, social stigma discouraging individuals from disclosing EVD exposure, and community mistrust^8,9,10^. To increase coverage, responders shifted to targeted geographic vaccination (TGV), offering vaccination to entire communities near a case regardless of known contact status^7^.

These experiences reveal a gap between the high efficacy of RV observed in trials and the more modest population-level effectiveness attainable under operationally-constrained field conditions, raising questions about circumstances under which TGV versus RV is most effective. Epidemiological and economic modelling can address these questions. However, previous modelling studies have mostly evaluated single strategies^11,12,13,14,15,16,17^, used stylised scenarios such as preventive versus reactive vaccination or retrospective RV in specific outbreaks –without directly comparing RV and TGV across a range of outbreak settings^18,19^, or assessed cost-effectiveness using simplified models unable to capture spatial heterogeneity or integration of contact tracing and response measures that characterise real outbreaks^20,21,22^. Consequently, existing models offer limited insight into the relative performance of RV and TGV under realistic conditions.

To address this gap, we developed a spatially-explicit individual-based stochastic EVD transmission model to directly compare reactive RV and TGV for Zaire species epidemics (hereafter we use EVD to mean EVD induced by *Orthoebolavirus zairense*). The model has informed WHO-led evidence synthesis and international discussions on Ebola vaccination and stockpiling strategies^23^. Using DRC as an exemplar setting with frequent outbreaks, we simulate a range of epidemic scenarios varying transmission intensity, detection delay, and response (e.g. contact tracing, case ascertainment, safe and dignified burials, treatment), and we compare outcomes for TGV and RV. Our aim is to inform policy on optimal reactive vaccination strategies and efficient use of vaccine supplies. This model does not use data from the 2026 Ebola outbreak caused by *Orthoebolavirus bundibugyoense*, against which no vaccines have been approved.

## 2 Methods

### 2.1 Model structure

Here we provide an overview of the EVD simulation model and the scenarios; for more detail, see the Supplementary Information (Section 1). The *ebolasim* microsimulation model generates a demographically-realistic synthetic population in North-East DRC capturing household, extended family, and community contact structures (Figure 1); simulates EVD transmission and disease progression within that population, including postmortem transmission; and implements outbreak response interventions including safe and dignified burials, contact tracing, specialised Ebola treatment centres, and vaccination.

**Figure 1:**
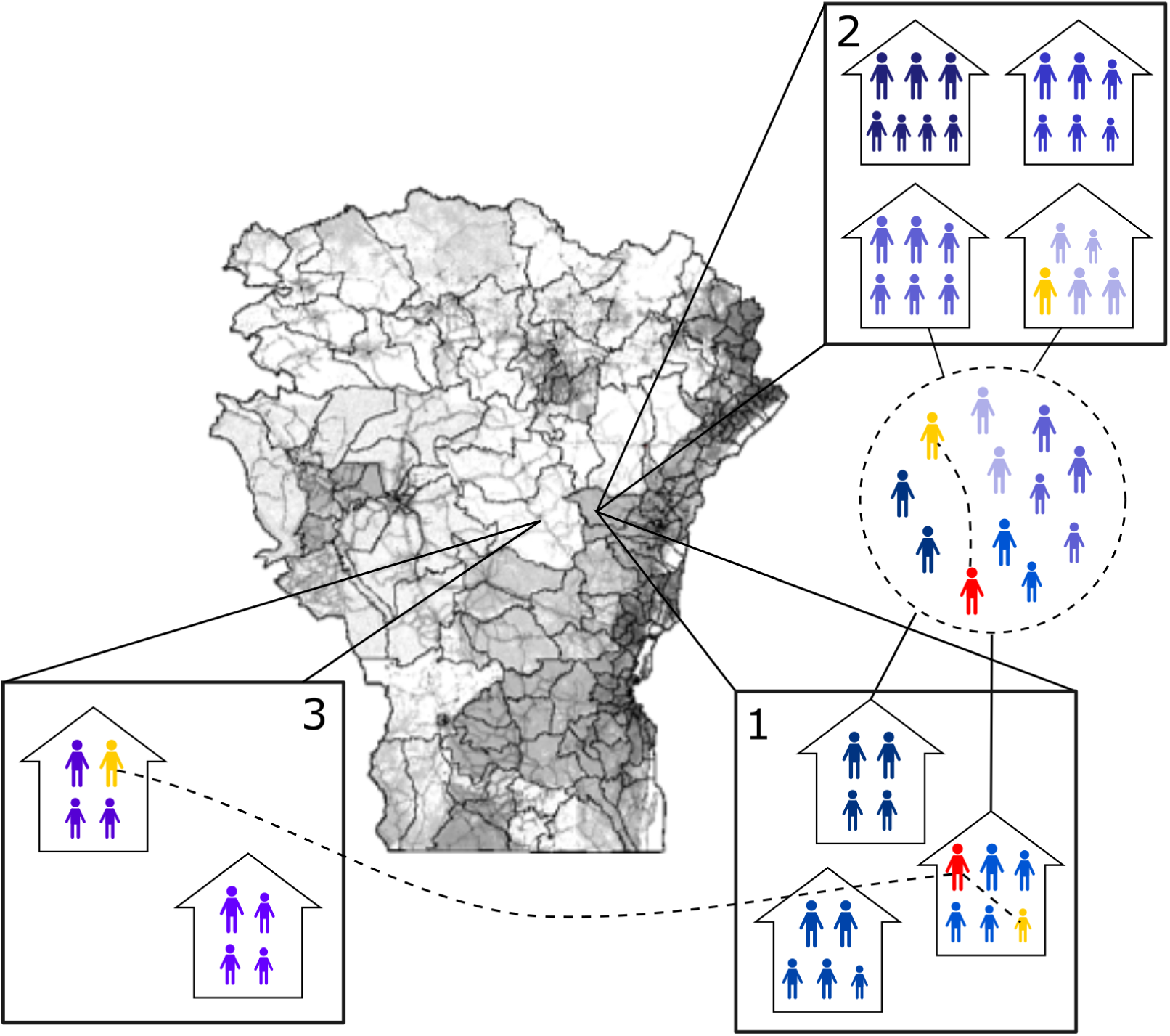
Illustrative schematic of the transmission model. The map shows the 39 territories of six Northeastern provinces of the Democratic Republic of the Congo included in the simulation, shaded by population density. Darker shading indicates higher density. The surrounding panels illustrate how the model represents space and contacts: grid cells divide the landscape; the population groups into households (coloured in different shades of blue) and extended family networks; transmission can occur within households, within extended families, and the wider community. Each cell contains fixed households of sizes sampled from demographic data. In this illustration, an infectious individual (red) infects a now-exposed household member (yellow) in cell 1. The dotted circle represents an extended family network mixing household members with neighbouring households in the same or adjacent cells. The infectious individual also infects an extended family member via that network, as well as a nearby community member to whom they do not have a household or family link, as shown in cell 3.

The community contact network reflects empirical observations from previous EVD RV campaigns in DRC^7^ and transmission and disease progression are represented using empirically-informed parameters from previous outbreaks^24^. Individual infectiousness varies between cases to capture transmission heterogeneity and potential super-spreading events^25^. The period of increased postmortem infectiousness represents transmission associated with funeral practices, and is calibrated to observed funeral infection events during the 2014-2016 West African outbreak^25^.

In the model, outbreak response activities are triggered after routine surveillance detects the first case. An Ebola Treatment Centre (ETC) is established in a territory (administrative level 2) once five cases have been detected there. Isolation in ETCs reduces an individual’s contacts by 80%, following assumptions that isolation greatly reduces transmission but not completely**^Shen^**,^26^. All fatal hospitalised cases and 80% of fatal non-hospitalised but detected cases receive safe and dignified burials^27,28^, reducing postmortem infectiousness probability by 80%^25^. Contact tracing teams are deployed after case detection and a proportion of the case’s household members and extended family network are followed-up for 21 days^29^, with a proportion lost to incomplete follow-up. Vaccination (either RV or TGV) begins seven days after outbreak detection^30^ and protects against EVD after 10 days^5^ with 84% effectiveness^5^. Under RV, a two-level ring of contacts and contacts-of-contacts is established with scenario-based probability (Table 1) with 95% vaccine acceptance within the rings^7^. Under TGV, vaccines are administered to individuals in households nearby to a case^7^, with scenario-based vaccine uptake probability (Table 1). For TGV, cases in an area triggers deployment of doses to that area; RV instead triggers for each detected case. Both strategies are subject to the same daily vaccine administration limits.

**Table 1:** Simulation model parameters for worst-case, moderate-case, and best-case outbreak responses. In each response scenario, the probability for TGV uptake is equal to the total probability of RV (which is the probability of establishing a ring multiplied by the probability of accepting vaccination given the ring establishment).

| Parameters |  | Response scenario |  |  | Ref |
| --- | --- | --- | --- | --- | --- |
| Parameter type | Parameter name | *Worst-case* | *Moderate-case* | *Best-case* |  |
| Case ascertainment | Proportion of ascertained cases who are not traced contacts | 0.33 | 0.61 | 0.83 | <sup>35</sup> |
|  | Fixed delay from symptom onset to case ascertainment (days) | 8 | 6 | 3 | <sup>7</sup> |
| Contact tracing | Proportion of contacts traced | 0.74 | 0.85 | 0.96 | Expert consultation |
|  | Proportion of contacts lost to follow-up | 0.50 | 0.25 | 0.05 | <sup>36,27</sup> |
| Vaccination | Fixed delay from symptom onset to vaccination triggered (days) | 10 | 7 | 3.5 | <sup>4</sup> |
|  | Probability of establishing a ring for RV | 0.66 | 0.83 | 1 | <sup>7</sup> |
|  | Vaccine uptake for RV | 0.95 | 0.95 | 0.95 | <sup>7</sup> |
|  | Vaccine uptake for TGV | 0.63 | 0.78 | 0.95 | Expert consultation |

### 2.2 Scenarios and parameterisation

We consider three transmission levels (low (*R*_0_=1.5), medium (*R*_0_=1.8), and high (*R*_0_=2.1), spanning the range of published estimates for EVD^31^), three outbreak detection times (early (20 undetected infections before outbreak detection), medium (40), and late (100), which reflect previous estimates^32,33^), three main response scenarios (reasonable worst-case, moderate-case, and best-case parameter sets for case ascertainment, contact tracing, and vaccination), and two sensitivity response scenarios, resulting in a total of 45 scenarios (Table 1). The parameters were chosen from available literature and where unavailable, via expert consultation. For sensitivity response scenarios, we allow the effectiveness of TGV to be greater than the rest of the non-vaccination activities by pairing best-case TGV parameters with (i) worst-case case ascertainment and contact tracing, and (ii) moderate-case ascertainment and contact tracing and compare them to the worst-case and moderate-case RV responses, respectively. This is to account for perceived acceptability of TGV compared to RV, due to the high level of stigma attached to being an EVD case contact^10^, therefore TGV may be carried out efficiently even when case ascertaining and contact tracing is difficult.

We simulate EVD outbreaks by seeding a single infected individual in Nord Kivu. We perform 500 stochastic outbreak simulations for each scenario and output the daily numbers of infections, detected cases, deaths, safe and dignified burials, vaccine doses used, contacts traced, and outbreak length, out to a maximum four year horizon. To estimate total vaccine dose requirements, we use the 95^th^ percentile (P95) of the simulated dose distributions as a proxy, reflecting the need for decision makers to plan for the tail risk of large outbreak scenarios.

We calculate outcomes related to health impacts and costs, including deaths and years of life lost (YLL) averted (Supplementary section 1.5). To compare vaccination strategy cost-effectiveness, simulations are paired using identical random number generator seeds so that epidemics evolve identically until vaccination is introduced. We calculate the incremental net monetary benefit (INMB) of TGV relative to RV using outbreak response cost estimates from the 2018-2020 Nord Kivu EVD outbreak^34^ (Supplementary section 1.5, Table S1). To understand whether differences in costs or net benefits drive the shape of the INMB distributions, we visualise incremental benefits and costs of TGV compared to RV on cost-effectiveness planes.

### 2.3 Ethics statement

This study did not involve human participants or identifiable data. It is a modelling study based on simulated populations and previously published data. Accordingly, it does not constitute human subjects research, and institutional review board/research ethics committee approval was not required. This activity was reviewed by the Centers for Disease Control and Prevention (CDC), determined to be non-research, and conducted consistent with applicable federal law and CDC policy.

## 3 Results

We present the moderate transmission level results (*R*_0_=1.8), with results for low (*R*_0_=1.5) and high (*R*_0_=2.1) transmission scenarios presented in Supplementary Section 2.

### Epidemiological impact

Tables S2-S5 and Figure 2 compare simulated total cases, deaths, and epidemic length across worst-, moderate-, and best-case response scenarios for TGV and RV strategies. Overall, no strategy consistently outperforms the other across all scenarios; their relative performance depends on the outbreak context, with differences between strategies consistently smaller than differences in outbreak detection timing and response effectiveness.

**Figure 2:**
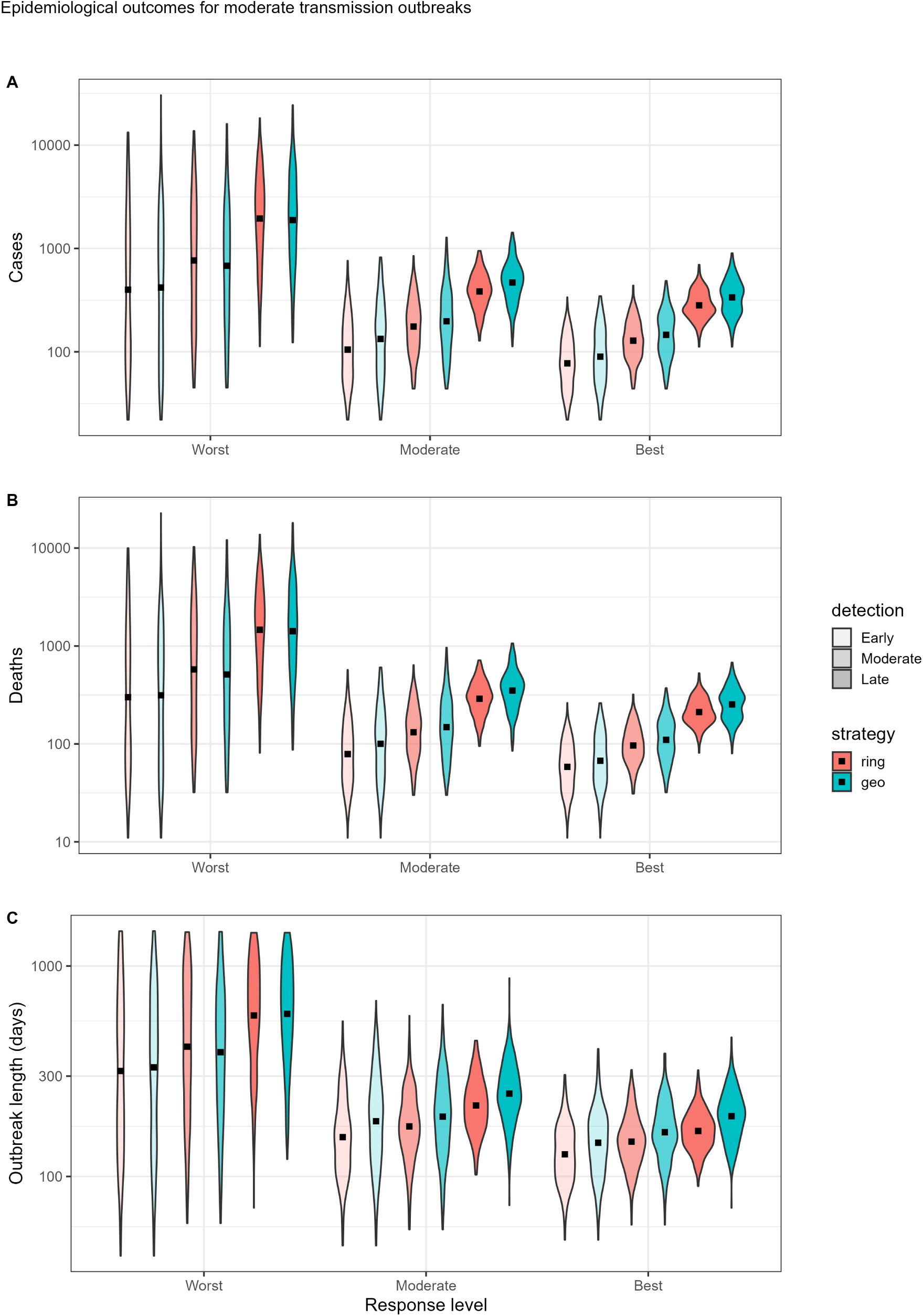
Violin plots showing the distributions of A: projected number of cases, B: projected number of deaths, and C: projected epidemic length (in days) for worst-case (left), moderate-case (centre), and best-case (right) outbreak responses (see Table 1) when employing ring (blue) and targeted geographic (red) vaccination strategies, and for early, medium, and late response times (indicated by transparency levels). Results are for the moderate transmission level scenario (*R*_0_=1.8). Note that the y-axis of all panels is shown on a log scale. The mean of each distribution is marked with a black square.

Under moderate– and best-case response scenarios, RV generally outperforms TGV at reducing mortality and epidemic size and duration. For example, under a moderate-case response scenario with medium outbreak detection time, the mean outbreak size is 18% smaller under RV: 210 cases (standard error on the mean [SEM]=8, 90% prediction interval [90%PI]: 58-468) compared to 256 cases (SEM=12, 90%PI: 59-660) under TGV. Corresponding average mortality outcomes are 158 deaths (SEM=6; 90%PI: 46-350) under RV compared to 192 (SEM=9; 90%PI: 46-486) under TGV. Differences between the strategies become larger when outbreak detection is delayed; under late detection the mean outbreak sizes increase to 415 (SEM=10, 90%PI: 186-740) cases and 520 (SEM=15, 90%PI: 217-1041) for RV and TGV, respectively, a difference of 105 cases (20%). Corresponding mortality outcomes are 313 (SEM=8; 90%PI: 146-548) versus 390 (SEM=11; 90%PI: 164-786) deaths. If TGV is delivered more effectively than underlying case ascertainment and contact tracing, RV retains a 14% advantage on average: best-case TGV conditions paired with moderate-case overall outbreak response gives a mean epidemic size of 243 (90% CI: 59-535).

In contrast, under worst-case response scenarios, TGV tends to outperform RV. Worst-case response scenarios exhibit substantially greater variability in epidemic outcomes, with large differences between outbreak sizes. Differences between strategies are most apparent in the upper ranges of the outbreak distributions where there are fewer very large outbreaks under TGV. For example, under medium outbreak detection, mean outbreak size is 13% greater with 1812 cases (90%PI: 70-7245) under RV compared to 1585 (90%PI: 66-5829) under TGV (Table S2). The modest advantage observed for TGV under worst-case response conditions becomes more pronounced when worst-case overall outbreak response is paired with TGV implemented under best-case vaccination conditions, corresponding to mean outbreak size 1,437 cases (90%CI: 65-6,104), 21% lower than under worst-case RV.

Patterns in the relative impact of RV and TGV are qualitatively similar across alternative transmission scenarios (Figures S4-S5) with RV performing better than TGV in moderate– and best-case response scenarios, although the magnitude of differences varies with transmission intensity. Under low transmission, RV and TGV strategies have broadly similar impact when outbreaks are detected early (Figure S4), and as outbreak detection is delayed, RV generally maintains a slight advantage (Tables S6-S8). Under high transmission, the marginal advantage of RV over TGV is more pronounced under moderate– and best-case outbreak response conditions, where RV results in larger reductions in epidemic size and duration. For example, under high transmission with moderate response, mean outbreak size is around 30% lower under RV (603 cases, 90%PI:98-1551) compared to TGV (898 cases, 90%PI:97-2508). Interpretation of worst-case response scenarios under high transmission is more difficult because many outbreaks continue to grow throughout the four-year simulation period, generating prolonged epidemics with very large numbers of cases and deaths under both vaccination strategies. Response limitations generate large intervention backlogs, preventing vaccination and case ascertainment efforts from keeping pace with ongoing transmission and newly emerging hotspots. This pattern suggests that, under the assumed response constraints, neither vaccination strategy is sufficient to control transmission, although RV performs marginally better.

While our central comparison is between RV and TGV, it is important to note that broader outbreak response has a greater influence on outcomes than the choice of vaccination strategy itself. Under a moderate outbreak response scenario with a medium detection time and TGV, the matched comparison with RV reduces the mean outbreak size by 46 cases (18%) from 256 (SEM=12, 90%PI: 59-660) to 210 (SEM=8; 90%PI: 58-468). In comparison, moving to the best-case response scenario reduced the mean outbreak size by 90 cases (35%) to 166 (SEM=5, 90%PI: 61-331), while earlier outbreak detection reduced mean outbreak size by 68 cases (27%) to 188 (SEM=9, 90%PI: 33-571). These findings suggest that improvements in outbreak detection and response capacity may have a larger influence on epidemic outcomes than optimisation of vaccination strategy alone.

### Vaccine dose requirements

We estimate the vaccine doses sufficient to respond to 95% of simulated outbreaks. Dose requirements differ between vaccination strategies and response scenarios (Table 2). Generally, the vaccination strategy with greater epidemiological impact tends to require fewer doses, since more effective control of transmission reduces outbreak growth and consequently lowers vaccine demand. Under moderate– and best-case response scenarios, requirements remain below 110,000 doses regardless of strategy, but RV consistently requires fewer doses than TGV. For example, under moderate response conditions with medium outbreak detection delay, mean vaccine requirement is reduced by 26% from 30,200 doses (90%PI: 2,700-91,400) under TGV to 22,400 doses (90%PI: 2,000-57,400) under RV. Worst-case response scenarios require substantially more doses and show the opposite pattern, for example, dose requirements reach a mean of 143,600 (90%PI: 2,100-579,700) under RV, which falls by 21% to 113,100 (90%PI: 2,600-378,800) under TGV. Similar patterns are observed across alternative transmission scenarios (Tables S9,S13). Under low transmission, dose requirements remain comparatively modest: RV requires fewer doses across all responses including worst-case, remaining below 61,000 doses, compared to 67,000 for TGV. Under high transmission, dose requirements increase but remain below approximately 280,000 doses for both RV and TGV for moderate– and best-case response conditions. Under worst-case response, estimated dose requirements increase substantially – up to 1.5 million for RV, and 690,000 for TGV; however, these estimates should be interpreted cautiously as outbreaks remain uncontrolled in many simulated worst-case scenarios, so true dose demand could be higher.

**Table 2:** Mean, 50^th^, and 95^th^ percentiles of the number of vaccine doses administered for ring (RV) and targeted geographic (TGV) vaccination strategies, for worst-case, moderate-case, and best-case response levels, and early, medium, and late outbreak detection times, rounded to the nearest 100 doses. Shaded cells indicate which vaccination strategy uses the fewest doses for the given scenario and summary statistic.

| Detection time | Summary statistic | Worst-case |  | Moderate-case |  | Best-case |  |
| --- | --- | --- | --- | --- | --- | --- | --- |
|  |  | RV | TGV | RV | TGV | RV | TGV |
| Early | Mean | 109,500 | 87,700 | 15,100 | 23,600 | 11,200 | 15,700 |
|  | 50<sup>th</sup> | 30,400 | 43,200 | 10,100 | 16,600 | 8,200 | 12,200 |
|  | 95<sup>th</sup> | 523,800 | 298,300 | 47,300 | 72,600 | 29,000 | 42,000 |
| Medium | Mean | 143,600 | 113,100 | 22,400 | 30,200 | 17,000 | 22,300 |
|  | 50<sup>th</sup> | 66,700 | 63,300 | 17,800 | 23,600 | 14,000 | 19,000 |
|  | 95<sup>th</sup> | 579,900 | 378,800 | 57,400 | 91,400 | 39,500 | 52,500 |
| Late | Mean | 235,400 | 196,800 | 42,500 | 53,400 | 32,600 | 39,500 |
|  | 50<sup>th</sup> | 175,400 | 165,600 | 38,500 | 51,000 | 28,900 | 38,800 |
|  | 95<sup>th</sup> | 676,200 | 477,300 | 87,400 | 109,500 | 60,900 | 70,700 |

### Cost-effectiveness

We calculate incremental net monetary benefits (INMB) using deaths averted as our measure of benefit for TGV relative to RV, to determine how the observed differences in outcomes between strategies affect cost-effectiveness (Figure 3). Across all scenarios, INMB distributions are wide and centred close to zero, indicating considerable stochastic variability in comparative cost-effectiveness. Similar patterns are observed under low and high transmission settings (Figures S7 and S9). The strategy that shortens the outbreak generally results in fewer cases and deaths, and reduced expenditure on costly outbreak response activities. The results are consistent when we use YLLs and discounted YLLs as alternative measurements of benefit (Figures S11-S22).

**Figure 3:**
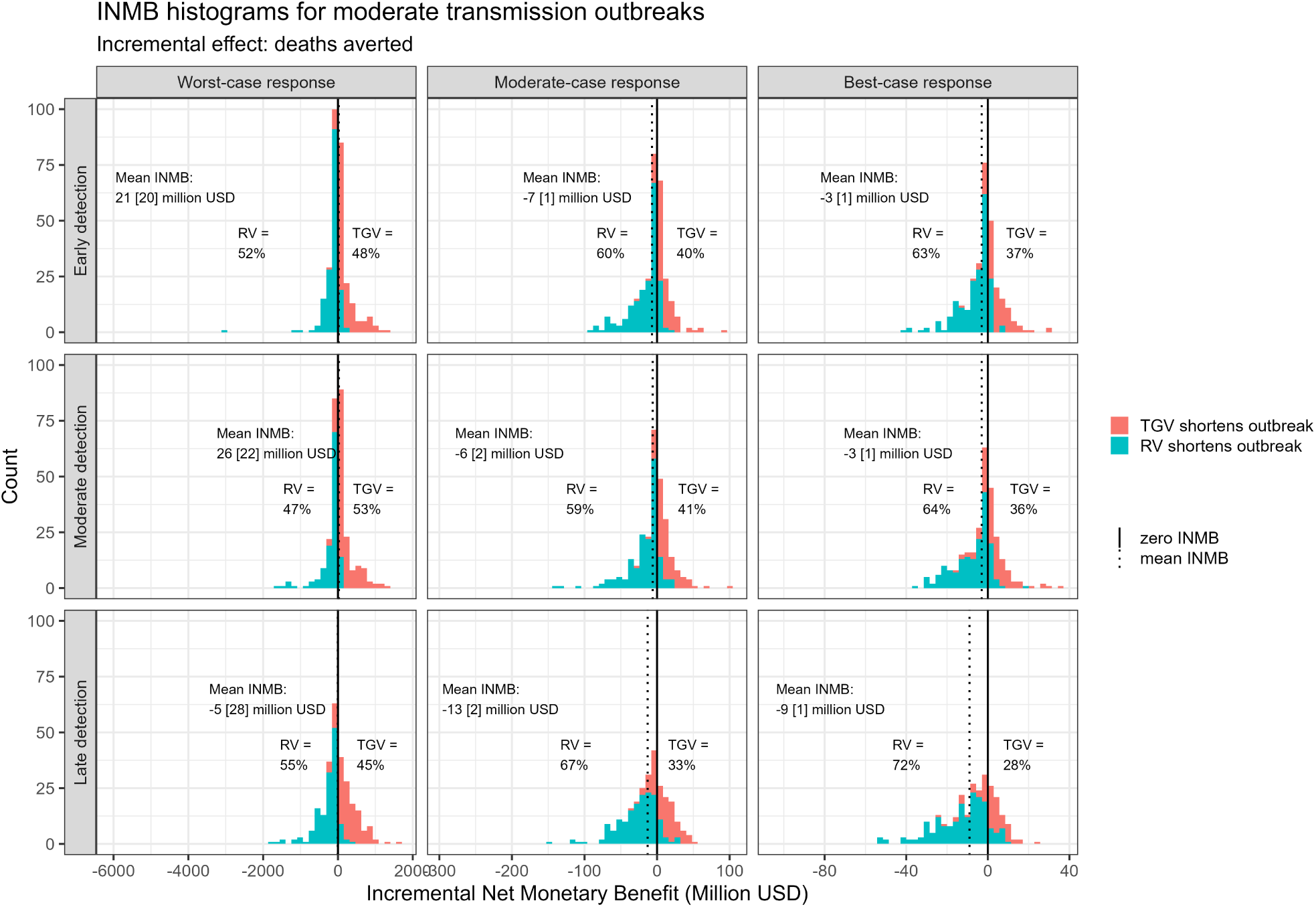
Histograms showing the incremental net monetary benefit (INMB) for TGV relative to RV. Detection time varies across the rows (top: early, middle: medium, bottom: late) and response level increases across the columns from left to right. Positive values of INMB (in red) indicate that TGV shortens outbreak more than RV, and negative values (in blue) indicate that RV results in shorter outbreaks. In each subplot, the mean INMB for the given scenario is shown, along with the standard error on the mean (SEM) in brackets, and the percentage of positive and negative INMBs.The histograms are colour-coded according to which strategy shortens outbreak length.

Consistent with the epidemiological findings, RV was slightly more likely to have greater benefits than TGV under moderate– and best-case response scenarios, as indicated by the greater proportion of negative INMBs (Figure 3). For example, RV has greater benefits in 59–67% of simulations under moderate-case responses, and 57-60% of simulations in the sensitivity analysis where moderate-case response was paired with best-case TGV. While TGV slightly outperformed RV under worst-case response scenarios in terms of epidemiological impact, the INMBs of strategies could not be differentiated, with TGV preferred in 48–53% of simulations. In the sensitivity analysis where worst-case response was paired with best-case TGV, TGV is preferred in 58-54% of simulations (Figure S6).

Under low transmission settings, differences between vaccination strategies are small, with INMB distributions tightly centred around zero and neither strategy consistently preferred (Figures S7 and S8). Under high transmission settings the relative advantage of RV becomes more pronounced, particularly under moderate– and best-case response scenarios where RV was preferred in 58–68% and 68–82% of simulations, respectively (Figures S9-S10). As discussed previously, estimates under high-transmission worst-case response scenarios should be interpreted cautiously, as continued epidemic growth may produce unstable cost-effectiveness comparisons.

## 4 Discussion

We evaluated the trade-offs of TGV compared to the recommended strategy of RV during *Orthoebolavirus zairense* EVD outbreaks, assuming a wide range of epidemiological scenarios and outbreak response constraints.

In moderate– or best-case outbreak response scenarios, RV on average produces better outcomes (fewer cases and deaths, shorter outbreaks, fewer vaccines required, and better cost-effectiveness) than TGV. This reflects the ability of RV to break transmission chains when case ascertainment is high and contact tracing swiftly identifies individuals to be vaccinated who are at highest risk of infection. By contrast, under worst-case conditions where contact tracing for RV is insufficient to break transmission chains, TGV offers a slight epidemiological advantage by establishing some community immunity and requires fewer doses. From an economic perspective, the results are equivocal for worst-case response scenarios, with each strategy dominating the other in roughly an equal number of simulations. This unpredictability means that decision-makers cannot confidently identify the more cost-effective vaccination strategy a priori under challenging outbreak conditions. Although we identify differences between RV and TGV strategies in terms of impact and cost-effectiveness, the mean differences are small and subject to considerable uncertainty. At the level of an individual outbreak, it is not possible to predict which strategy will perform better, even though consistent patterns emerge when results are averaged across many simulated outbreaks. We note that the timeliness of detection and efficacy of ascertainment, isolation, and response have a substantially greater impact than the choice of vaccination strategy itself.

Our analysis has limitations. We explicitly model safe and dignified burials as one component of Infection Prevention and Control (IPC); however, other components – such as case isolation practices, environmental decontamination, and linen and waste management in health-care facilities and communities^37^ – entail additional costs and benefits. Future work should incorporate a broader range of IPC, including Water, Sanitation, and Hygiene (WASH) measures in the response framework. Furthermore, our estimates of vaccine dose requirements do not account for wastage and we only consider one outbreak at a time when, in reality, multiple outbreaks may occur within a given period. Additionally, our cost-effectiveness analysis may not capture all relevant operational costs, such as one-off start-up costs, which disproportionately effect shorter outbreaks. We treat RV and TGV as distinct intervention strategies to enable a clear comparison. In practice, under challenging field conditions, RV may approximate TGV, as difficulties in contact tracing can lead teams to vaccinate households or communities surrounding known cases rather than strictly contacts^38^. Conversely, behavioural responses may cause TGV to resemble RV, for example, vaccine uptake in an area is likely to be non-random if vaccine acceptance increases with proximity to known cases^39^. In our comparisons, the impact of TGV in the absence of RV should therefore be interpreted as a lower bound, reflecting scenarios in which contact tracing has largely broken down. In practice, the effectiveness of TGV would likely be greater, as some RV would typically continue in parallel.

Another assumption concerns the fixed contact structure, where 82% of interactions are within households/extended families rather than the community (broadly consistent with observations during the West African outbreak^25^); by construction determining the proportion of the modelled population that can be traced. In settings characterised by higher population density or greater mobility, the share of community interactions may be higher, potentially altering the relative performance of RV versus TGV. For parameters based on West African EVD, if detailed data specific to DRC become available, our estimates can be refined. Data from DRC EVD in 2018-2020 informed the assumed degree of spatial clustering in the model, the vaccine delivery rates, and the contact tracing capacity. Greater spatial dispersion would likely affect both the impact of TGV and the feasibility of effective contact tracing for RV. During 2018-2020, vaccination was initialised under the compassionate-use protocol which required trained vaccinator teams, registration and consent procedures, and adverse effect followup periods. Vaccine delivery rates may be higher in the post-licensure context. Contact tracing capacity may be lower than in previous outbreaks (as is being observed in the current Bundibugyo ebolavirus (BVD) outbreak^40^, although no vaccines have been approved for this species), which would likely increase the preference for TGV. The current BVD outbreak highlights the need to continue revisiting and refining parameter assumptions.

Previous studies have evaluated combined ring and community vaccination^41^, hypothesised the potential benefits of TGV^42^, and retrospectively analysed the number of EVD cases that could have been averted using RV compared to other vaccination strategies during the 2018-2020 EVD outbreak in DRC^19^. Our work provides an important addition by directly comparing health outcomes and cost-effectiveness of vaccination strategies during simulated future EVD outbreaks across a full range of epidemiological and operational contexts. Our methods can be expanded to explore vaccination strategy prioritisation under different assumptions of vaccine effectiveness; for example, against BVD for which vaccine candidates are in development^43,44^. Furthermore, the cost-effectiveness of preventively vaccinating frontline and health-care workers could be explored.

Overall, our findings support RV – currently the recommended standard-of-care^45^ – as a highly effective strategy for *Orthoebolavirus zairense* EVD outbreaks. However, TGV should be considered a viable contingency option, particularly in settings where operational constraints limit the effectiveness of RV. This suggests that vaccination strategy can be chosen pragmatically based on outbreak context, without sacrificing impact. Perhaps the key finding of this work is that, while vaccination is an important addition to the outbreak response toolbox, it is not a panacea; to curtail EVD outbreaks, we need robust surveillance and outbreak response measures.

## Competing interests

K. Hauck and A. Cori received personal fees from Munich Re for work unrelated to this project. A. Bagayoko, A. Costa, and P. Lambach work for the World Health Organization (WHO). The authors alone are responsible for the views expressed in this publication and they do not necessarily represent the decisions, policy, or views of the WHO. The findings and conclusions in this report are those of the authors and do not necessarily represent the views of the U.S. Centers for Disease Control and Prevention.

## Funding

GNG, VMC, JES, RJ, WH, CM, RM, LKW, NMF, KH, and AC acknowledge funding from the MRC Centre for Global Infectious Disease Analysis (reference MR/X020258/1), funded by the UK Medical Research Council (MRC). This UK funded award is carried out in the frame of the Global Health EDCTP3 Joint Undertaking. GNG, VMC, JES, RJ, WH, CM, RM, LKW, NMF, and AC acknowledge joint funding from the UK MRC and the UK Foreign, Commonwealth and Development Office (FCDO) under the MRC/FCDO Concordat agreement (grant number UKRI2029). NMF and KH acknowledge funding by Community Jameel. CABP contributed to this project via cooperative agreement CDC-RFA-FT-23-0069 from the CDC Center for Forecasting and Outbreak Analytics. Its contents are solely the responsibility of the authors and do not necessarily represent the official views of the Centers for Disease Control and Prevention. SR is currently seconded to UKHSA as Chief Data Officer. This work was supported by Gavi, the Vaccine Alliance. The authors alone are responsible for the views expressed in this publication and they do not necessarily represent the decisions, policy or views of Gavi.

## Code availability

The code and data required to run and reproduce the analysis presented here is available at https://github.com/mrc-ide/ebolasim_public.

## Supporting information

Supplementary methods and results

## Data Availability

The code used to produce these results is available at https://github.com/mrc-ide/ebolasim_public

## Notes

### Competing Interest Statement

KH and AC received personal fees from Munich Re for work unrelated to this project.

### Summary of Updates

Revised the text to reduce the word count, moved more of the methods to supplementary material, revised the figures, added additional authors who have contributed to the manuscript.

