## Supplementary methods and results for "Cost-effectiveness of targeted geographic versus ring vaccination campaigns in response to an outbreak of Ebola Virus Disease caused by *Orthoebolavirus zairense*"

July 2026

### 1 Supplementary Methods

Here we describe the individual-based model used to simulate EVD outbreaks, the model parameters, and the range of scenarios explored in this study. The *ebolasim* microsimulation model consists of three linked components: a population model, a disease transmission model, and an intervention and response model. The underlying framework used to generate geographically-distributed synthetic populations and simulate outbreaks builds on established spatial microsimulation methods that have previously been applied across a range of infectious diseases [1]. We provide full methodological details for each component below.

#### 1.1 Population model

We used 2018 Landscan data for the Democratic Republic of the Congo (DRC) [2] and GADM shape files [3] to generate our synthetic population and allocate them to spatial cells within level 2 administrative units (territories). We simulate the population of North-East DRC focusing on 39 territories that comprise six provinces (Nord Kivu, Ituri, Haut-Uele, Tshopo, Maniema and Sud Kivu), with a combined population of 22.3 million people. The model divides the area into a high-resolution grid of spatial cells approximately 1km$^2$ in size. The population distribution within each spatial cell is informed by regional demographic data [2]. Individuals within each spatial cell are assigned ages and grouped into households according to demographic data published following the 2013-2014 demographic and health survey carried out in DRC [4]. Figures S1 and S2 show the input and output distributions for age and household size respectively. Note that during the household set-up phase of the simulation, ages within the household are adjusted according to heuristic rules [1] to preserve intergenerational age differences. This results in the age distribution of the synthetic population diverging from the input distribution. Specifically, we see a slight reduction in the under-15 age bands. At present we do not include age-specific infection and mortality rates in the model as the extent to which observed age differences in EVD burden reflect intrinsic biological differences versus differences in exposure, contact patterns, and healthcare-seeking behaviour remains uncertain. Although incorporation of age structure could affect quantitative estimates, we would not expect the modest shifts in population age structure observed here to substantially alter conclusions regarding the relative epidemiological impact of vaccination strategies. However, in the cost-effectiveness analysis, benefits are measured by the number of discounted years life lost averted. Skewing the population slightly towards older age group could lead to a modest underestimation of intervention benefit. Future work will focus on refining the household generation procedure to better capture the underlying population age structure and incorporating age-dependent effects where evidence is stronger, such as age-specific mortality rates [5].

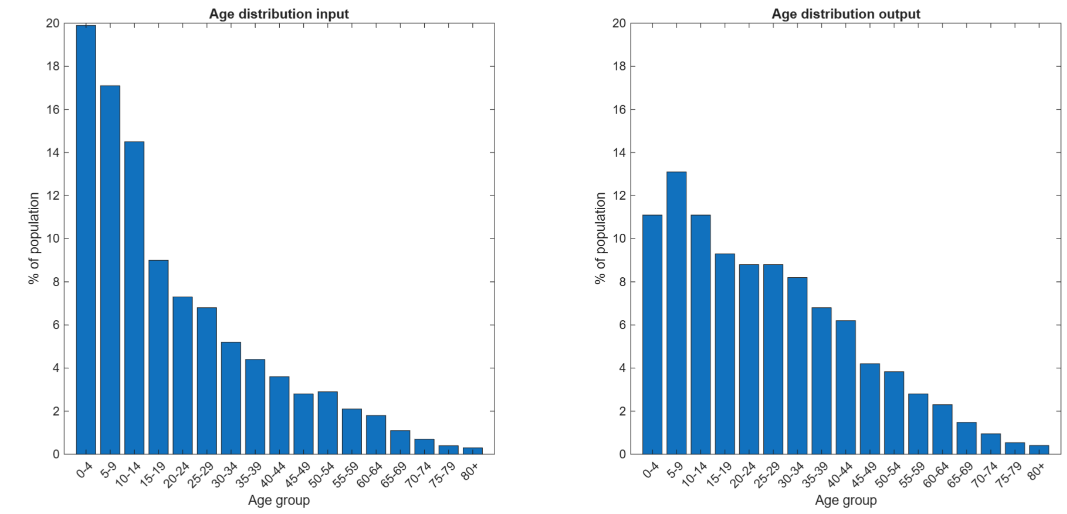


Figure S1: Left: target population age distribution from [4] and right: synthetic population age distribution.

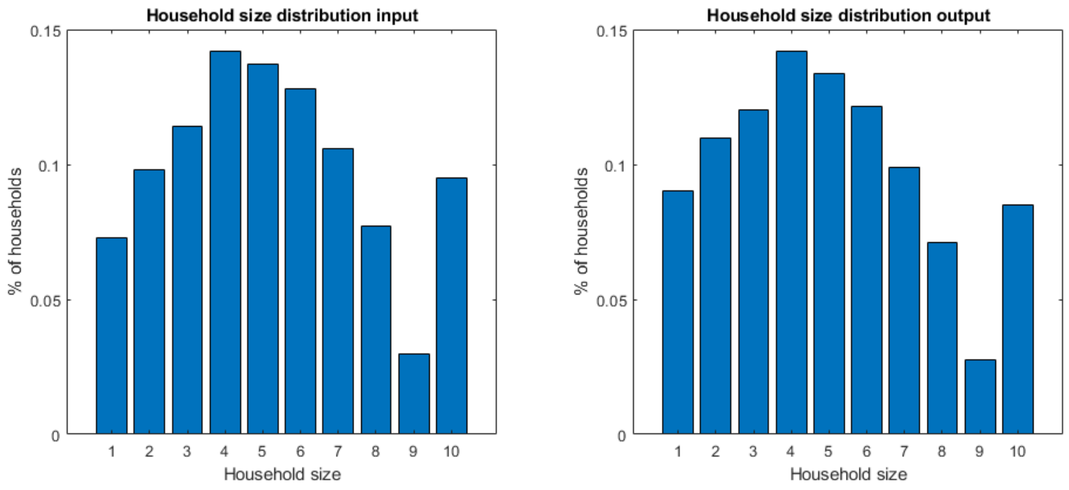


Figure S2: Left: target population household size distribution from [4] and right: synthetic population household size distribution.

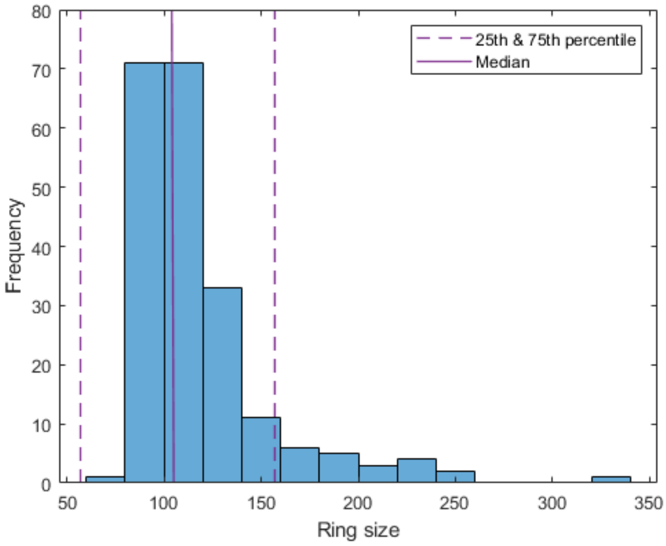


Figure S3: Vaccination ring size frequency taken from a simulated population whose extended family network sizes are drawn from a log normal distribution with $\mu = 40$ and $\sigma = 40$. The purple lines indicate the median and interquartile range of vaccination ring sizes in [6].

Individuals are then allocated to an extended family group which, along with household members, forms the list of contacts (and contacts-of-contacts) used for contract tracing and ring vaccination. The sizes of extended family groups are drawn from a log normal distribution with mean $\mu$ and standard deviation $\sigma$. We assume that there is some overlap between household and extended family membership; when looping over individuals and assigning them to an extended family group, we also assign roughly half of their household members to the same extended family group. Whilst they do not represent explicit locations, the availability of extended family groups depends on population density and individuals are preferentially allocated to one of the three groups closest to them. Therefore individuals in areas of high population density are more likely to share an extended group with individuals in the same spatial cell whereas individuals in areas of low population density are more likely to share an extended family group with individuals from other spatial cells. To calibrate the size distribution of the extended family group, we varied $\mu$ and $\sigma$ and compared the size of the vaccination rings generated by the households and extended family networks to data on vaccination ring size [6]. We found $\mu = 40$ and $\sigma = 40$ most closely matched the mean ring size from the data.

#### 1.2 Transmission model and disease natural history parameters

Under the transmission model, the force of infection, $\lambda_i$, on an individual $i$, is given by

$$\lambda_i = \sum_{k|h_k=h_i} \frac{I_k \beta_h \rho_k}{n_i^{\alpha}} + \sum_{k|f_k=f_i} \frac{I_k \beta_f \rho_k}{m_i} + \frac{\sum_k I_k \beta_c \rho_k f(d_{i,k})}{\sum_k f(d_{i,k})}, \tag{1}$$

where the first term represents household force of infection, the second term represents extended family network force of infection, and the third term captures the community force of infection. $k$ only contributes to the household force of infection if $i$ and $k$ share the same household, i.e. $h_k = h_i$, and to the extended family force of infection if they share an extended family network, i.e. $f_k = f_i$. $I_k = 1$ if an individual is infectious and 0 otherwise, $\beta_h$, $\beta_f$, and $\beta_c$ are the respective household, extended family, and transmission coefficients, $\rho_k$ is individual $k$'s infectiousness, $n_i$ is the number of members in $i$'s household, $\alpha$ is a scaling factor [7], $m_i$ is the number of members in $i$'s extended family network, and $f(d_{i,k})$ is the value of the spatial kernel at distance $d_{i,k}$ between individuals $i$ and $k$.

We calibrate the transmission coefficients assuming a baseline $R_0$ value of 1.8, such that households account for 42% of transmission events, extended family networks account for 40% of transmission events, and community transmission making up the remaining 18% of infections, roughly matching contact patterns observed during the West African Ebola outbreak [8]. This results in transmission coefficients of $\beta_h = 0.088/day$, $\beta_f = 0.033/day$, and $\beta_c = 0.029/day$. Note that when we consider lower or higher transmission settings, these $\beta$ values are scaled accordingly. In the absence of data describing the relationship between household size and household attack rate specifically for EVD, we use the value derived by Cauchemez *et al* for influenza [7], i.e $\alpha = 0.8$. $\rho_k$ is drawn from a gamma distribution with mean 1 and standard deviation 2 to match offspring distribution data from the West African Ebola outbreak [8], as described in the main text.

When an individual is infected, we sample their incubation period from a gamma distribution with a mean of 10.2 days and a standard deviation of 6.0 days [8]. The individual becomes infectious after the incubation period. The length of their infectious period is sampled from a Weibull distribution with scale parameter 10.6 and a shape parameter 1.42 [9]. The individual's outcome is determined by comparing the relative probability of recovery or death, given the infectious period duration, as described in the main text. When an individual who will go on to die reaches the end of their infectious period, we simulate post-mortem transmission by extending their infectious period for an another day and increasing their infectiousness by a factor of 3.4, which allows us to match the proportion of funeral exposures recorded in [8].

The spatial kernel is a gravity model in the form of a power-law function, as in [1], and is described by

$$f(d_{i,k}) = \frac{1}{1 + (\frac{d_{i,k}}{a})^b}. \tag{2}$$

Estimating the parameters of such a model for Sub-Saharan African countries is challenging as most available data sources focus on long-term migration trends rather than on short-term movement. To try and understand the spatial dynamics of the 2018-2020 North Kivu outbreak, we consider the distance between different generations of cases using publicly available data (https://data.humdata.org/dataset/ebola-cases-and-deaths-drc-north-kivu). Using the spatial and temporal distances between the first ever recorded case and the first recorded cases in other territories and using a serial interval of 15.3 days [9], we estimate the the virus travels a distance of 6-20km per generation. We choose a spatial kernel with a=1km and b=1.2, for which there is a 75% probability that the distance travelled is below 20km whilst still allowing a small probability for long distance transmission events. However, we note that without specific data to support this, the choice of kernel parameters is slightly arbitrary. Many different combinations of parameters will also produce a similar spatial profile but may result in slightly different spatial dynamics.

##### 1.3 Outbreak detection and response

For each realisation in the simulation, we seed the infection of one randomly-chosen individual in Nord Kivu. Following the end of the incubation period, the individual becomes infectious and on the next time step develops symptoms. Their infectiousness remains constant until they recover or die. The first detection of an EVD case through routine, passive surveillance triggers outbreak response activities. We consider three initial outbreak detection scenarios: early, intermediate, or late. These scenarios are characterised by the number of missed infections occurring prior to the first detection of a case. The number of initial undetected infections can range from tens [10] to hundreds [11, 12]. Following outbreak detection, a proportion of cases are ascertained, depending on whether the outbreak response level matches the worst-case, moderate-case, or best-case as described in Table 1. Outbreaks that go extinct before being detected, and therefore do not prompt an outbreak response, are not included in the analysis.

For outbreak response and Infection Prevention and Control (IPC), we denote the time of first detection of a case as time zero, against which delays in rolling out and duration of interventions are measured. Following outbreak detection, an Ebola Treatment Centre (ETC) is established in each territory (administration level 2) once five cases have been detected there. We assume a two-week construction period, after which the ETC becomes operational with a capacity of 12 beds. When the ETC is at 80% capacity, it is earmarked for expansion; after a delay of seven days, an additional 12 beds become available. This process can be repeated multiple times, dependent on occupancy, and each time the expansion takes seven days and results in an additional 12 beds. However, a new expansion cannot begin until the previous one has been completed. We assume a fixed delay from case detection to hospitalisation, which is reduced for individuals who are contacts under active investigation. If a bed is available in the ETC of a case's residential territory, they are admitted until recovery or death, which reduces their contacts with household, family, and community members by 80% through isolation practices; otherwise, they remain in the community. Safe and dignified burial teams are deployed seven days after outbreak detection [13]. We assume that all fatal hospitalised cases and 80% of fatal non-hospitalised but detected cases are given safe and dignified burials [14, 15] and that a safe and dignified burial reduces the postmortem infectiousness probability of the deceased individual by 80% [8].

Contact tracing teams are activated two days after outbreak detection [16], and deployed to each territory once a case has been detected there. When a detected case triggers contact tracing, a proportion of their household members and extended family network are added to a list of contacts to follow for up to 21 days [17], of which a proportion are lost to follow-up. We assume that each territory has the capacity to follow up 5000 contacts at any given time, based on the maximum reported number of contacts traced per active health zone [18] multiplied by the average number of health zones per territory. If the territory is at contact tracing capacity when a case is detected, the contacts of the case are not added to the contact tracing list, even if capacity were to become available later on in the 21-day period following detection of the case. A proportion of contacts being traced are lost to follow-up during the 21-day period, and the day on which they are lost to follow-up is randomly selected. We assume that traced contacts who become cases during their follow-up period are detected on the day of symptom onset.

Vaccination roll-out, in either a ring or targeted geographic strategy, begins seven days after the outbreak is detected [16]. We assume a delay of 10 days for vaccinated individuals to be protected by the vaccine [19], after which the vaccine is 84% effective [19]. We assume that the vaccine does not reduce the infectiousness or severity of breakthrough cases and that effectiveness does not wane over the simulation period.

At the beginning of the response, vaccination teams can vaccinate a maximum of 500 individuals per day across all affected regions; this is increased to 1000 individuals per day when the average number of cases ascertained exceeds one per day over the past seven days on average, and to a maximum of 1500 individuals per day when the average number of cases ascertained per day exceeds two per day over the past seven days and then remains at this level for the rest of the simulation. If vaccination demand exceeds capacity, individuals are added to a vaccination 'queue' to be vaccinated on a subsequent day, and they remain in the queue until they have been vaccinated or become ineligible (due to symptom onset). Individuals are added to the queue according to when they were identified as eligible for vaccination and the queue is processed in order using random tie-breaks where necessary.

Under a targeted geographic strategy, a vaccination team is deployed to a case's $1km^2$ grid cell with 300 doses of vaccine per case to administer to individuals living nearby [6], up to a maximum of 900 doses (personal communication, Dr Aminata Bagayoko, January 2024). We randomly sample households and vaccinate all household members aged 12 months or over [20], subject to assumed vaccine uptake probability (Table 1) until all the available vaccine doses have been used. We assume that all household members either all accept, or all reject vaccination. If the number of residents in the cell (the spatial grid unit of the model) where vaccination is being rolled-out is less than the number of doses available, the vaccination teams start vaccinating households in neighbouring cells. They start in the closest cells and move out to more distant cells until either all doses have been used or up to a maximum distance of 5km from the ascertained case's cell.

#### 1.4 Ebola Virus Disease outbreak response costs

To provide cost estimates for each scenario we accounted for the cost of contact tracing, surveillance, and laboratory tests (per simulated ascertained case), total cost of treatment which is the cost of case management in both treatment and transit centres (per simulated hospitalised case), cost of vaccination (per vaccine given), cost of safe burial (per safe burials conducted), and the monthly cost of the rapid response team and control surveillance per territory (for the length of the simulated outbreak). The outbreak length in months is defined as the time from the first to the last ascertained case, plus 42 days to account for the active observational period following the final case [21], divided by 30 days per month. We use unit costs where possible; however, we calculate the unit cost per safe and dignified burial by dividing the budgeted costs for safe and dignified burials [22] by the 22,921 safe and dignified burials listed in the situation reports by the end of the 2018-2020 North Kivu outbreak [23, 24]. We calculated the total cost of the response by summing the costs of all interventions.

To estimate the outbreak response cost of a given realisation, we match the unit costs calculated by Zeng et al [22] to outputs from the simulation model. These costs and the matched outputs are given in table S1. We do not include the psychosocial support costs as we do not explicitly include frontline workers, who are beneficiaries of this, in this version of the model.

| Activity | Unit cost (USD) | Simulation output unit |
|---|---|---|
| Rapid response team (per team per month) | 66,182 | Number of months |
| Point of entry (per point of entry per month) | 18,293 | Admin unit months |
| Community engagement (per capita per month) | 2.71 | Population affected months |
| Contact tracing (per identified case) | 4,435 | Number of detected cases |
| Laboratory testing (per identified case) | 4,166 | Number of detected cases |
| Treatment cost (per confirmed case) | 1,683 | Number of hospitalised cases |
| Vaccination (per vaccinated individual) | 120.70 | Number of vaccinations |
| Safe burials (per burial) | 1,758 | Number of safe burials |

Table S1: Unit costs taken from [22] to estimate outbreak response cost. Note that the cost of case management in treatment and transit centres is summed to give a single treatment cost and that the unit cost of a safe and dignified burial is approximated from the budget given in [22] and the number of safe and dignified burials given in [23, 24]

#### 1.5 Health economics analysis

To compare the cost-effectiveness of the vaccination strategies, simulations are paired using identical random number generator seeds so that epidemics evolve identically until vaccination is introduced. We then calculate the incremental net monetary benefit (INMB) of TGV relative to RV as

$$\mathrm{INMB}_i^j = \mathrm{VSL}(\mathrm{D}_{ring,i}^j - \mathrm{D}_{geo,i}^j) - (\mathrm{C}_{geo,i}^j - \mathrm{C}_{ring,i}^j), \tag{3}$$

where $\mathrm{D}_{geo,i}^j$ and $D_{ring,i}^j$ are the number of deaths for realisation $i$ and scenario $j$ associated with TGV and RV respectively, $\mathrm{C}_{geo,i}^j$ and $\mathrm{C}_{ring,i}^j$ are the corresponding outbreak costs, and $\mathrm{VSL}$ is the value of a statistical life (VSL). Cost estimates are based on data from the 2018-2020 Nord Kivu EVD outbreak [22] and include vaccination, surveillance, contact tracing, laboratory testing, treatment, safe and dignified burials, and other outbreak response activities. Full costing assumptions are provided in Supplementary Table S1. We use $\mathrm{VSL} = \$107,000$ as the average VSL for a lower income country [25]. We use this instead of a DRC-specific VSL as we believe that non-governmental organisations and international aid agencies paying for vaccination campaigns are likely to use a single, representative value when comparing the cost-effectiveness of different interventions across different countries with similar income levels.

For comparison of our results with cost-effectiveness studies using alternative metrics, we also present $\mathrm{INMB}^{\mathrm{YLL}}$ calculated using years of life lost $(\mathrm{YLL})$ as

$$\mathrm{INMB}_i^{\mathrm{YLL},j} = \mathrm{VSLY}(\mathrm{YLL}_{ring,i}^j - \mathrm{YLL}_{geo,i}^j) - (\mathrm{C}_{geo,i}^j - \mathrm{C}_{ring,i}^j), \tag{4}$$

where $\mathrm{YLL}_{geo,i}^j$ and $\mathrm{YLL}_{ring,i}^j$ now represent the years of lives lost for realisation $i$ and scenario $j$ associated with TGV and RV respectively, and $\mathrm{VSLY}$ is the value of a statistical life year. Assuming that $\mathrm{VSL}$ is a weighted average of remaining life years multiplied by a constant $\mathrm{VSLY}$ [26], we calculate $\mathrm{VSLY}$ using health-adjusted life expectancy figures for DRC [27] and our previously chosen $\mathrm{VSL}$, resulting in an estimated $\mathrm{VSLY}$ of $2877.

We also present $\mathrm{INMB}^{\mathrm{DYLL}}$ calculated using discounted $\mathrm{YLL}$ $(\mathrm{DYLL})$ as

$$\mathrm{INMB}_i^{\mathrm{DYLL},j} = \sum_y \left( \left( \mathrm{VSLY}(\mathrm{YLL}_{ring,i}^{y,j} - \mathrm{YLL}_{geo,i}^{y,j}) - (\mathrm{C}_{geo,i}^{y,j} - \mathrm{C}_{ring,i}^{y,j}) \right) (1+d)^{-y} \right), \quad (5)$$

where $\mathrm{YLL}$s and costs are discounted at rate $d$ according to the year $y$ in which they occur and then summed over all years of the outbreak. We use an equal discount rate of $d = 0.03$ for both $\mathrm{YLL}$ and $\mathrm{C}$.

### 2 Supplementary Results

In this section, we provide additional results for the moderate transmission scenario ($R_0$=1.8) and full results for the low transmission ($R_0$=1.5) and high transmission ($R_0$=2.1) settings.

#### 2.1 Epidemiological Impact

##### 2.1.1 Moderate transmission settings

Tables S2 to S5 show summary statistics of the case, death, epidemic duration, and vaccine dose distributions for moderate transmission settings.

| | | Response Level | | | | | | | |
|---|---|---|---|---|---|---|---|---|---|
| | | Worst-case | | | Moderate-case | | | Best-case | |
| Detection Time | Summary Stat | Ring | Geo | Best Geo | Ring | Geo | Best Geo | Ring | Geo |
| Early | Mean | 1377 | 1182 | 994 | 139 | 188 | 157 | 90 | 109 |
| | 5th | 36 | 36 | 36 | 33 | 33 | 33 | 31 | 32 |
| | 25th | 98 | 111 | 121 | 56 | 66 | 63 | 48 | 56 |
| | 50th | 394 | 510 | 376 | 105 | 146 | 115 | 78 | 88 |
| | 75th | 1573 | 1431 | 1283 | 184 | 258 | 206 | 119 | 150 |
| | 95th | 6397 | 3767 | 3971 | 358 | 561 | 421 | 190 | 245 |
| Medium | Mean | 1812 | 1585 | 1437 | 210 | 256 | 243 | 142 | 166 |
| | 5th | 71 | 67 | 65 | 60 | 59 | 59 | 59 | 62 |
| | 25th | 212 | 239 | 210 | 112 | 114 | 122 | 94 | 100 |
| | 50th | 818 | 716 | 605 | 184 | 197 | 200 | 128 | 150 |
| | 75th | 2354 | 1966 | 1613 | 272 | 339 | 322 | 178 | 221 |
| | 95th | 7243 | 5606 | 6104 | 464 | 650 | 535 | 266 | 330 |
| Late | Mean | 3048 | 3122 | 2571 | 415 | 520 | 497 | 296 | 362 |
| | 5th | 345 | 288 | 302 | 187 | 218 | 199 | 178 | 182 |
| | 25th | 996 | 864 | 734 | 291 | 344 | 309 | 230 | 250 |
| | 50th | 2296 | 2026 | 1547 | 396 | 483 | 454 | 271 | 348 |
| | 75th | 4044 | 4348 | 3392 | 510 | 631 | 611 | 355 | 438 |
| | 95th | 8599 | 8710 | 7101 | 732 | 1027 | 970 | 470 | 610 |

Table S2: Summary statistics for the distribution of number of cases in a moderate transmission scenario.

| | | Response Level | | | | | | | |
|---|---|---|---|---|---|---|---|---|---|
| | | Worst-case | | | Moderate-case | | | Best-case | |
| Detection Time | Summary Stat | Ring | Geo | Best Geo | Ring | Geo | Best Geo | Ring | Geo |
| Early | Mean | 1034 | 888 | 748 | 104 | 142 | 118 | 68 | 82 |
| | 5th | 27 | 27 | 27 | 26 | 26 | 26 | 24 | 25 |
| | 25th | 72 | 80 | 89 | 44 | 51 | 49 | 38 | 42 |
| | 50th | 287 | 390 | 280 | 80 | 111 | 84 | 58 | 66 |
| | 75th | 1180 | 1070 | 956 | 138 | 190 | 153 | 90 | 112 |
| | 95th | 4823 | 2822 | 2976 | 264 | 431 | 315 | 142 | 185 |
| Medium | Mean | 1362 | 1191 | 1081 | 158 | 192 | 182 | 106 | 125 |
| | 5th | 55 | 52 | 52 | 47 | 47 | 45 | 47 | 47 |
| | 25th | 166 | 177 | 157 | 87 | 84 | 92 | 72 | 76 |
| | 50th | 617 | 540 | 456 | 138 | 148 | 150 | 95 | 112 |
| | 75th | 1761 | 1447 | 1205 | 204 | 252 | 241 | 133 | 162 |
| | 95th | 5420 | 4217 | 4611 | 349 | 482 | 406 | 196 | 257 |
| Late | Mean | 2292 | 2345 | 1933 | 313 | 390 | 373 | 222 | 273 |
| | 5th | 253 | 210 | 231 | 146 | 165 | 150 | 131 | 139 |
| | 25th | 744 | 652 | 556 | 218 | 258 | 232 | 170 | 188 |
| | 50th | 1719 | 1556 | 1159 | 297 | 361 | 334 | 207 | 263 |
| | 75th | 3071 | 3262 | 2552 | 386 | 472 | 454 | 264 | 326 |
| | 95th | 6397 | 6539 | 5358 | 546 | 777 | 731 | 353 | 457 |

Table S3: Summary statistics for the distribution of number of deaths in a moderate transmission scenario.

| | | Response Level | | | | | | | |
|---|---|---|---|---|---|---|---|---|---|
| | | Worst-case | | | Moderate-case | | | Best-case | |
| Detection Time | Summary Stat | Ring | Geo | Best Geo | Ring | Geo | Best Geo | Ring | Geo |
| Early | Mean | 454 | 451 | 428 | 170 | 207 | 186 | 134 | 155 |
| | 5th | 87 | 82 | 82 | 82 | 81 | 79 | 77 | 78 |
| | 25th | 156 | 156 | 180 | 112 | 128 | 122 | 99 | 109 |
| | 50th | 309 | 361 | 338 | 152 | 190 | 166 | 128 | 147 |
| | 75th | 679 | 681 | 604 | 208 | 266 | 228 | 164 | 187 |
| | 95th | 1355 | 1153 | 1068 | 316 | 419 | 339 | 214 | 257 |
| Medium | Mean | 542 | 500 | 462 | 186 | 212 | 211 | 153 | 171 |
| | 5th | 109 | 104 | 96 | 92 | 90 | 87 | 91 | 95 |
| | 25th | 222 | 227 | 204 | 138 | 142 | 148 | 121 | 134 |
| | 50th | 432 | 415 | 373 | 173 | 196 | 199 | 145 | 161 |
| | 75th | 831 | 690 | 614 | 224 | 269 | 252 | 173 | 203 |
| | 95th | 1267 | 1288 | 1157 | 301 | 366 | 358 | 245 | 266 |
| Late | Mean | 682 | 682 | 608 | 227 | 262 | 250 | 169 | 201 |
| | 5th | 194 | 203 | 202 | 136 | 142 | 135 | 113 | 124 |
| | 25th | 409 | 414 | 345 | 176 | 204 | 182 | 142 | 162 |
| | 50th | 634 | 632 | 535 | 215 | 249 | 245 | 165 | 196 |
| | 75th | 934 | 910 | 828 | 268 | 309 | 294 | 189 | 234 |
| | 95th | 1374 | 1301 | 1202 | 356 | 399 | 411 | 231 | 296 |

Table S4: Summary statistics for the distribution of epidemic length (in days) in a moderate transmission scenario.

| | | Response Level | | | | | | | |
|---|---|---|---|---|---|---|---|---|---|
| | | Worst-case | | | Moderate-case | | | Best-case | |
| Detection Time | Summary Stat | Ring | Geo | Best Geo | Ring | Geo | Best Geo | Ring | Geo |
| Early | Mean | 109530 | 87744 | 88136 | 15135 | 23648 | 20942 | 11165 | 15717 |
| | 5th | 1092 | 1300 | 1514 | 1102 | 1645 | 1898 | 1186 | 1926 |
| | 25th | 5899 | 7739 | 9729 | 3772 | 6386 | 6015 | 3640 | 5403 |
| | 50th | 30407 | 43182 | 41080 | 10137 | 16603 | 13678 | 8156 | 12226 |
| | 75th | 133519 | 129308 | 129174 | 21476 | 31186 | 29817 | 15349 | 22800 |
| | 95th | 523486 | 294468 | 325121 | 46810 | 72279 | 59919 | 28910 | 41646 |
| Medium | Mean | 143572 | 113128 | 111789 | 22388 | 30225 | 30314 | 17010 | 22296 |
| | 5th | 2137 | 2715 | 2942 | 2057 | 2692 | 2941 | 2342 | 3658 |
| | 25th | 15035 | 18695 | 17709 | 8610 | 9746 | 11904 | 8004 | 10770 |
| | 50th | 66741 | 63343 | 61474 | 17768 | 23562 | 25943 | 13985 | 19037 |
| | 75th | 194417 | 164812 | 164493 | 31554 | 42205 | 42915 | 24643 | 31890 |
| | 95th | 577880 | 377684 | 408758 | 56381 | 91335 | 72548 | 39115 | 51795 |
| Late | Mean | 235447 | 196848 | 182931 | 42481 | 53397 | 52513 | 32647 | 39452 |
| | 5th | 18408 | 16169 | 22910 | 11839 | 16280 | 13455 | 12309 | 14591 |
| | 25th | 70645 | 70321 | 70395 | 24760 | 32571 | 31085 | 21388 | 26020 |
| | 50th | 175365 | 165597 | 139742 | 38495 | 50954 | 50568 | 28901 | 38764 |
| | 75th | 329032 | 299548 | 289616 | 56326 | 67258 | 68036 | 43088 | 50401 |
| | 95th | 675578 | 470703 | 449182 | 87232 | 107704 | 107004 | 60807 | 70659 |

Table S5: Summary statistics for the distribution of number of vaccine doses administered in a moderate transmission scenario.

##### 2.1.2 Low transmission settings

Figure S4 shows swarm plots comparing the distributions of cases, deaths and epidemic duration for worst-case, moderate-case, and best-case outbreak responses and for all outbreak detection times, assuming a low transmission setting, and tables S6 to S9 show summary statistics of the case, death, epidemic duration, and vaccine dose distributions.

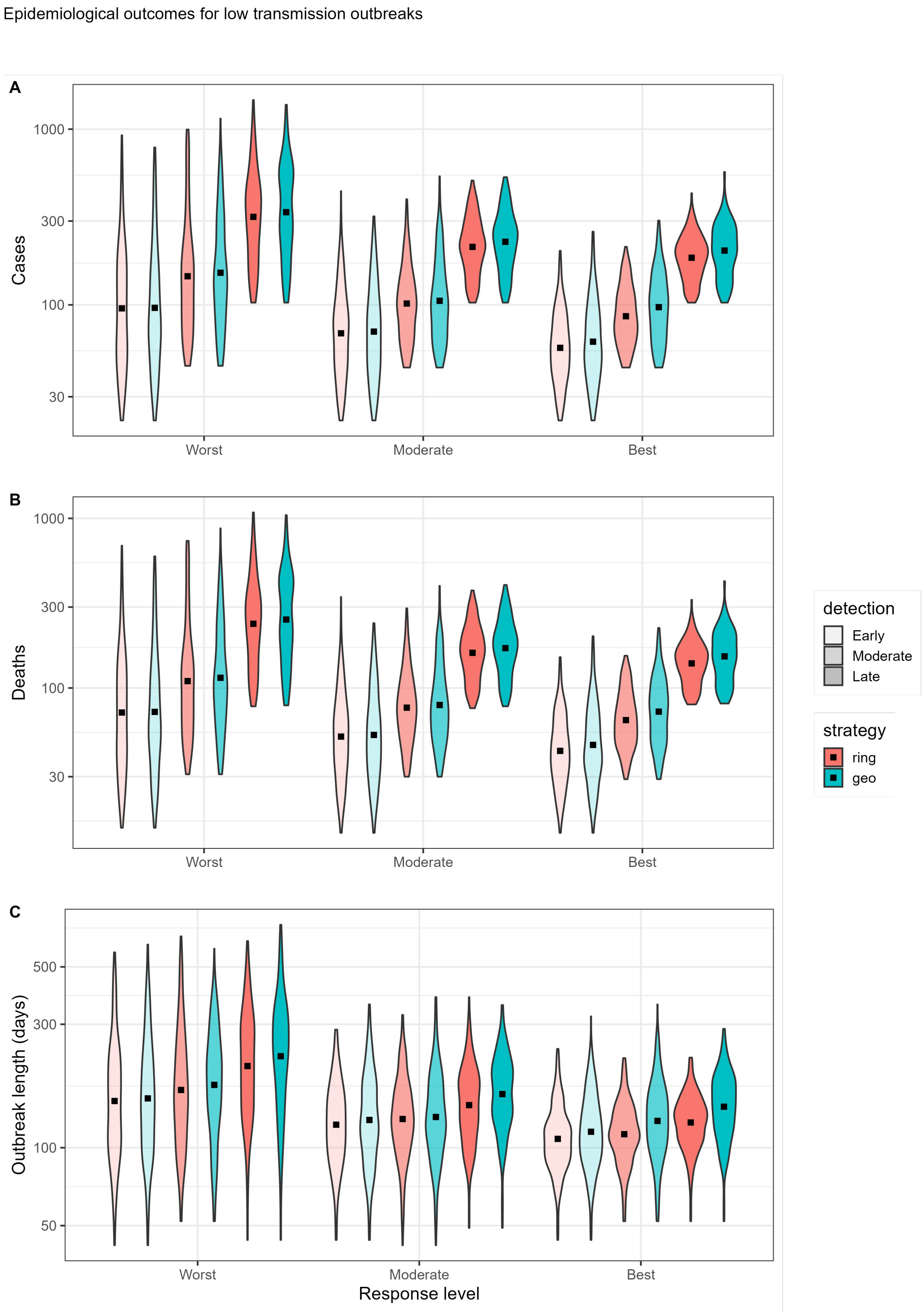


Figure S4: Violin plots showing the distribution of A: projected number of cases, B: projected number of deaths, and C: projected epidemic length (in days) for worst-case (left), moderate-case (centre), and best-case (right) outbreak responses (see main text for parameters) when employing ring (blue) and targeted geographic (red) vaccination strategies, and for early, medium, and late response times (indicated by transparency levels). Results are for the low transmission level scenario. Note that the y-axis of all panels is shown on a log scale. The mean of each distribution is marked with a black square.

| | | Response Level | | | | | | | |
|---|---|---|---|---|---|---|---|---|---|
| | | Worst-case | | | Moderate-case | | | Best-case | |
| Detection Time | Summary Stat | Ring | Geo | Best Geo | Ring | Geo | Best Geo | Ring | Geo |
| Early | Mean | 135 | 138 | 134 | 83 | 86 | 82 | 64 | 72 |
| | 5th | 30 | 30 | 30 | 28 | 28 | 28 | 27 | 28 |
| | 25th | 52 | 53 | 49 | 44 | 44 | 44 | 41 | 42 |
| | 50th | 93 | 85 | 85 | 71 | 69 | 65 | 57 | 58 |
| | 75th | 173 | 160 | 164 | 100 | 108 | 102 | 81 | 88 |
| | 95th | 387 | 445 | 368 | 187 | 207 | 202 | 122 | 147 |
| Medium | Mean | 203 | 199 | 191 | 115 | 123 | 125 | 93 | 107 |
| | 5th | 52 | 50 | 53 | 47 | 47 | 47 | 47 | 47 |
| | 25th | 81 | 88 | 86 | 71 | 68 | 72 | 65 | 68 |
| | 50th | 136 | 142 | 138 | 100 | 100 | 106 | 83 | 96 |
| | 75th | 219 | 257 | 259 | 137 | 154 | 150 | 113 | 134 |
| | 95th | 712 | 527 | 488 | 231 | 274 | 282 | 163 | 204 |
| Late | Mean | 379 | 404 | 369 | 229 | 247 | 237 | 193 | 217 |
| | 5th | 121 | 116 | 116 | 116 | 117 | 115 | 112 | 115 |
| | 25th | 198 | 213 | 205 | 163 | 174 | 170 | 150 | 154 |
| | 50th | 323 | 329 | 311 | 218 | 236 | 227 | 191 | 207 |
| | 75th | 476 | 556 | 508 | 279 | 300 | 294 | 228 | 265 |
| | 95th | 850 | 851 | 764 | 401 | 435 | 406 | 300 | 330 |

Table S6: Summary statistics for the distribution of number of cases in a low transmission scenario.

| | | Response Level | | | | | | | |
|---|---|---|---|---|---|---|---|---|---|
| | | Worst-case | | | Moderate-case | | | Best-case | |
| Detection Time | Summary Stat | Ring | Geo | Best Geo | Ring | Geo | Best Geo | Ring | Geo |
| Early | Mean | 102 | 104 | 101 | 62 | 64 | 62 | 48 | 54 |
| | 5th | 21 | 22 | 21 | 21 | 21 | 21 | 20 | 20 |
| | 25th | 38 | 41 | 40 | 34 | 33 | 33 | 30 | 33 |
| | 50th | 71 | 63 | 67 | 52 | 50 | 48 | 43 | 44 |
| | 75th | 126 | 118 | 126 | 78 | 81 | 78 | 60 | 68 |
| | 95th | 294 | 329 | 281 | 142 | 158 | 153 | 89 | 109 |
| Medium | Mean | 153 | 150 | 143 | 87 | 92 | 94 | 70 | 80 |
| | 5th | 38 | 38 | 39 | 35 | 35 | 35 | 35 | 35 |
| | 25th | 62 | 68 | 65 | 54 | 53 | 56 | 47 | 50 |
| | 50th | 104 | 106 | 102 | 75 | 75 | 80 | 64 | 75 |
| | 75th | 164 | 190 | 196 | 104 | 115 | 109 | 86 | 103 |
| | 95th | 535 | 396 | 375 | 174 | 201 | 210 | 125 | 152 |
| Late | Mean | 286 | 303 | 276 | 173 | 185 | 179 | 145 | 163 |
| | 5th | 91 | 91 | 91 | 87 | 88 | 88 | 87 | 89 |
| | 25th | 148 | 158 | 149 | 124 | 132 | 128 | 113 | 114 |
| | 50th | 244 | 246 | 235 | 163 | 176 | 169 | 142 | 156 |
| | 75th | 358 | 429 | 380 | 209 | 226 | 218 | 170 | 206 |
| | 95th | 641 | 634 | 572 | 303 | 328 | 312 | 221 | 259 |

Table S7: Summary statistics for the distribution of number of deaths in a low transmission scenario.

| | | Response Level | | | | | | | |
|---|---|---|---|---|---|---|---|---|---|
| | | Worst-case | | | Moderate-case | | | Best-case | |
| Detection Time | Summary Stat | Ring | Geo | Best Geo | Ring | Geo | Best Geo | Ring | Geo |
| Early | Mean | 174 | 178 | 175 | 131 | 139 | 133 | 113 | 122 |
| | 5th | 67 | 70 | 70 | 69 | 68 | 68 | 69 | 69 |
| | 25th | 103 | 102 | 104 | 94 | 96 | 94 | 89 | 92 |
| | 50th | 153 | 151 | 153 | 125 | 126 | 120 | 107 | 113 |
| | 75th | 218 | 221 | 214 | 159 | 171 | 165 | 126 | 144 |
| | 95th | 398 | 374 | 369 | 221 | 248 | 223 | 182 | 196 |
| Medium | Mean | 193 | 196 | 195 | 138 | 142 | 146 | 117 | 135 |
| | 5th | 79 | 75 | 77 | 68 | 74 | 70 | 71 | 69 |
| | 25th | 114 | 131 | 121 | 104 | 104 | 108 | 96 | 102 |
| | 50th | 158 | 183 | 181 | 128 | 128 | 140 | 113 | 128 |
| | 75th | 238 | 249 | 249 | 163 | 175 | 181 | 136 | 159 |
| | 95th | 431 | 366 | 381 | 234 | 239 | 246 | 181 | 216 |
| Late | Mean | 232 | 255 | 234 | 155 | 170 | 165 | 129 | 150 |
| | 5th | 97 | 93 | 93 | 86 | 94 | 93 | 85 | 90 |
| | 25th | 143 | 162 | 160 | 116 | 131 | 124 | 108 | 119 |
| | 50th | 211 | 241 | 220 | 149 | 166 | 160 | 129 | 147 |
| | 75th | 298 | 328 | 298 | 186 | 204 | 192 | 148 | 180 |
| | 95th | 446 | 496 | 420 | 244 | 267 | 263 | 178 | 228 |

Table S8: Summary statistics for the distribution of epidemic length (in days) in a low transmission scenario.

| | | Response Level | | | | | | | |
|---|---|---|---|---|---|---|---|---|---|
| | | Worst-case | | | Moderate-case | | | Best-case | |
| Detection Time | Summary Stat | Ring | Geo | Best Geo | Ring | Geo | Best Geo | Ring | Geo |
| Early | Mean | 8663 | 9994 | 12196 | 7244 | 8634 | 8904 | 6124 | 8527 |
| | 5th | 705 | 889 | 883 | 745 | 1138 | 1153 | 808 | 1208 |
| | 25th | 1810 | 2414 | 2549 | 2122 | 2688 | 3008 | 2184 | 3072 |
| | 50th | 5331 | 5117 | 6645 | 5006 | 5788 | 5942 | 4248 | 5858 |
| | 75th | 11459 | 12274 | 14854 | 9577 | 12069 | 11248 | 8456 | 11748 |
| | 95th | 30732 | 36296 | 44641 | 20184 | 25820 | 26946 | 17615 | 23680 |
| Medium | Mean | 12536 | 13810 | 16108 | 9140 | 11538 | 12893 | 7704 | 11794 |
| | 5th | 765 | 890 | 1174 | 753 | 1156 | 1032 | 911 | 1207 |
| | 25th | 2809 | 4056 | 4300 | 3076 | 3791 | 4696 | 3267 | 5104 |
| | 50th | 6996 | 8670 | 10923 | 7279 | 8226 | 9608 | 5998 | 10172 |
| | 75th | 13478 | 18164 | 24735 | 12509 | 17070 | 16854 | 10567 | 17058 |
| | 95th | 51986 | 43527 | 50900 | 24439 | 31772 | 40128 | 19440 | 26945 |
| Late | Mean | 21670 | 26674 | 28328 | 16530 | 20346 | 20308 | 14900 | 19388 |
| | 5th | 1376 | 1479 | 1810 | 1836 | 2915 | 2699 | 1998 | 2712 |
| | 25th | 7408 | 8648 | 9975 | 7146 | 10171 | 10246 | 7696 | 10549 |
| | 50th | 16927 | 19792 | 22361 | 14387 | 19016 | 19697 | 13972 | 19094 |
| | 75th | 29012 | 39934 | 40828 | 23427 | 28362 | 27461 | 20698 | 26258 |
| | 95th | 59975 | 66534 | 70409 | 37874 | 42091 | 46646 | 32581 | 39118 |

Table S9: Summary statistics for the distribution of number of vaccine doses administered in a low transmission scenario.

##### 2.1.3 High transmission settings

Figure S5 shows swarm plots comparing the distributions of cases, deaths, and epidemic duration for worst-case, moderate-case, and best-case outbreak responses and for all outbreak detection times, assuming a high transmission setting, and tables S10 to S13 show summary statistics of the case, death, epidemic duration, and vaccine dose distributions.

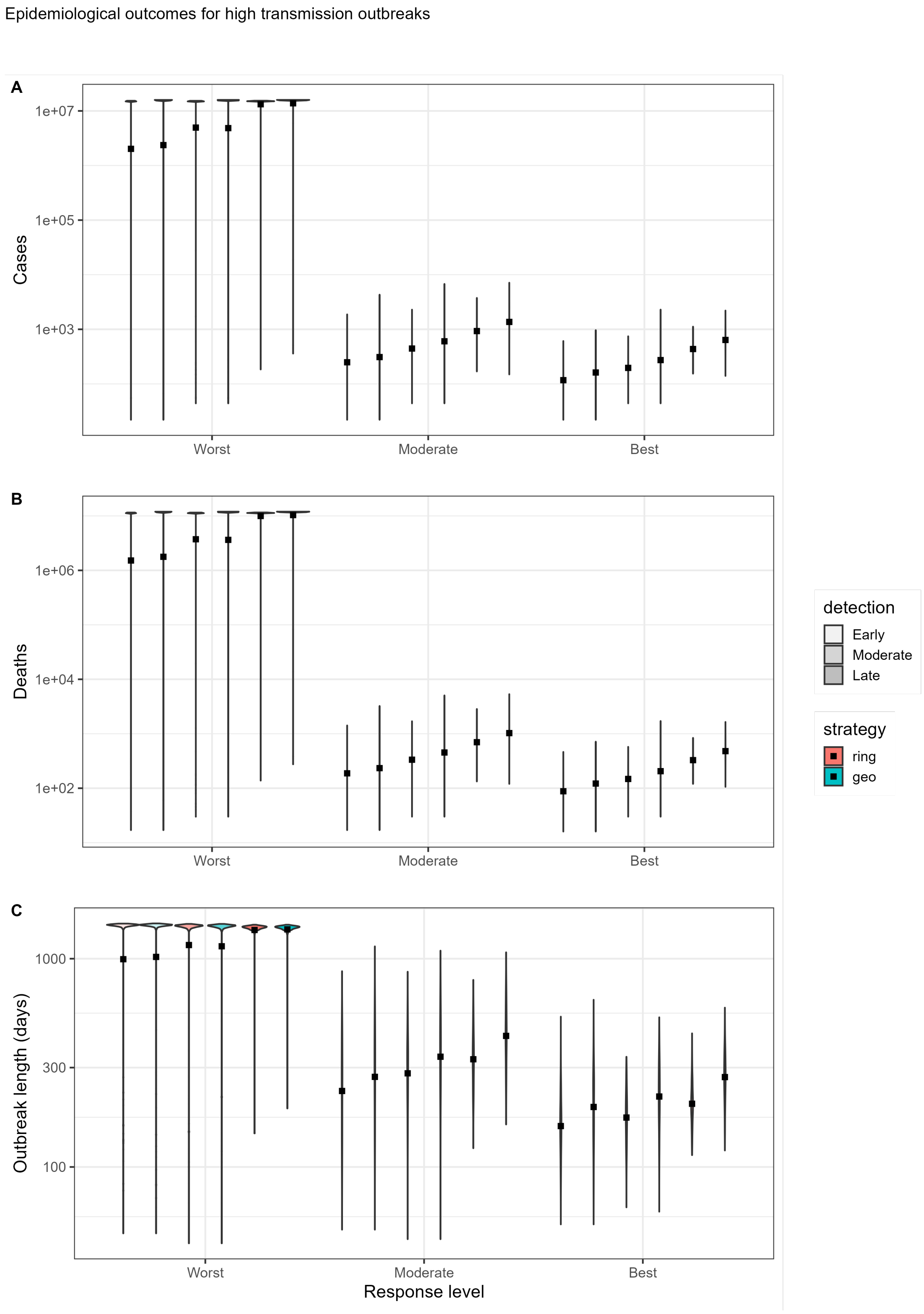


Figure S5: Violin plots showing the distributions of A: projected number of cases, B: projected number of deaths, and C: projected epidemic length (in days) for worst-case (left), moderate-case (centre), and best-case (right) outbreak responses (see main text for parameters) when employing ring (blue) and targeted geographic (red) vaccination strategies, and for early, medium, and late response times (indicated by transparency levels).Results are for the high transmission level scenario. Note that the y-axis of all panels is shown on a log scale. The mean of each distribution is marked with a black square.

| | | Response Level | | | | | | | |
|---|---|---|---|---|---|---|---|---|---|
| | | Worst-case | | | Moderate-case | | | Best-case | |
| Detection Time | Summary Stat | Ring | Geo | Best Geo | Ring | Geo | Best Geo | Ring | Geo |
| Early | Mean | 12558490 | 13147071 | 12430941 | 412 | 552 | 519 | 146 | 221 |
| | 5th | 55 | 53 | 43 | 39 | 44 | 41 | 35 | 37 |
| | 25th | 14980744 | 15685780 | 15587736 | 110 | 134 | 123 | 72 | 91 |
| | 50th | 15093469 | 15747188 | 15733507 | 270 | 314 | 305 | 120 | 168 |
| | 75th | 15168882 | 15798594 | 15791193 | 614 | 781 | 748 | 203 | 290 |
| | 95th | 15252008 | 15842054 | 15844725 | 1286 | 1837 | 1645 | 340 | 540 |
| Medium | Mean | 13659153 | 14058136 | 13866978 | 603 | 898 | 804 | 227 | 338 |
| | 5th | 128 | 111 | 154 | 99 | 99 | 92 | 75 | 80 |
| | 25th | 15037829 | 15712600 | 15696766 | 259 | 323 | 279 | 135 | 179 |
| | 50th | 15112998 | 15764162 | 15758192 | 470 | 684 | 564 | 206 | 288 |
| | 75th | 15175652 | 15806498 | 15803355 | 803 | 1210 | 1035 | 297 | 458 |
| | 95th | 15253963 | 15848529 | 15850023 | 1536 | 2425 | 2467 | 446 | 722 |
| Late | Mean | 14956230 | 15522732 | 15629242 | 1069 | 1688 | 1615 | 465 | 711 |
| | 5th | 14971899 | 15692750 | 15696865 | 329 | 416 | 424 | 228 | 275 |
| | 25th | 15090770 | 15769542 | 15767161 | 655 | 869 | 852 | 344 | 469 |
| | 50th | 15151908 | 15810541 | 15807731 | 960 | 1438 | 1346 | 448 | 665 |
| | 75th | 15196357 | 15843150 | 15838351 | 1363 | 2331 | 2140 | 575 | 880 |
| | 95th | 15262483 | 15879031 | 15884179 | 2146 | 3712 | 3662 | 754 | 1302 |

Table S10: Summary statistics for the distribution of number of cases in a high transmission scenario.

| | | Response Level | | | | | | | |
|---|---|---|---|---|---|---|---|---|---|
| | | Worst-case | | | Moderate-case | | | Best-case | |
| Detection Time | Summary Stat | Ring | Geo | Best Geo | Ring | Geo | Best Geo | Ring | Geo |
| Early | Mean | 9443232 | 9878767 | 9337871 | 310 | 415 | 390 | 110 | 167 |
| | 5th | 37 | 38 | 31 | 28 | 32 | 31 | 27 | 28 |
| | 25th | 11265300 | 11794200 | 11703900 | 84 | 101 | 92 | 54 | 68 |
| | 50th | 11350700 | 11843800 | 11834300 | 203 | 238 | 226 | 90 | 126 |
| | 75th | 11409450 | 11882600 | 11876500 | 459 | 592 | 554 | 154 | 217 |
| | 95th | 11470960 | 11915510 | 11916910 | 945 | 1373 | 1243 | 259 | 408 |
| Medium | Mean | 10272145 | 10567367 | 10415640 | 453 | 674 | 605 | 171 | 254 |
| | 5th | 97 | 84 | 117 | 71 | 76 | 65 | 57 | 57 |
| | 25th | 11309900 | 11818125 | 11805800 | 198 | 237 | 210 | 102 | 132 |
| | 50th | 11366200 | 11856000 | 11850950 | 361 | 509 | 428 | 157 | 215 |
| | 75th | 11413975 | 11889600 | 11885275 | 604 | 904 | 773 | 226 | 354 |
| | 95th | 11472750 | 11919375 | 11920385 | 1148 | 1818 | 1864 | 337 | 532 |
| Late | Mean | 11248485 | 11670978 | 11749191 | 805 | 1270 | 1216 | 350 | 534 |
| | 5th | 11261100 | 11799940 | 11804640 | 249 | 310 | 316 | 172 | 199 |
| | 25th | 11350000 | 11860400 | 11858400 | 496 | 640 | 649 | 258 | 353 |
| | 50th | 11395800 | 11890200 | 11889200 | 722 | 1082 | 1018 | 336 | 498 |
| | 75th | 11428900 | 11915500 | 11912000 | 1024 | 1744 | 1619 | 432 | 662 |
| | 95th | 11479300 | 11943320 | 11945280 | 1603 | 2753 | 2741 | 567 | 990 |

Table S11: Summary statistics for the distribution of number of deaths in a high transmission scenario.

| | | Response Level | | | | | | | |
|---|---|---|---|---|---|---|---|---|---|
| | | Worst-case | | | Moderate-case | | | Best-case | |
| Detection Time | Summary Stat | Ring | Geo | Best Geo | Ring | Geo | Best Geo | Ring | Geo |
| Early | Mean | 1236 | 1252 | 1203 | 268 | 317 | 304 | 167 | 213 |
| | 5th | 121 | 110 | 97 | 88 | 99 | 95 | 87 | 86 |
| | 25th | 1426 | 1428 | 1424 | 150 | 178 | 159 | 126 | 143 |
| | 50th | 1448 | 1448 | 1446 | 249 | 286 | 259 | 161 | 204 |
| | 75th | 1458 | 1457 | 1457 | 357 | 414 | 410 | 196 | 261 |
| | 95th | 1466 | 1466 | 1466 | 517 | 642 | 632 | 267 | 380 |
| Medium | Mean | 1316 | 1308 | 1304 | 311 | 384 | 349 | 181 | 234 |
| | 5th | 150 | 140 | 196 | 123 | 127 | 115 | 102 | 115 |
| | 25th | 1413 | 1413 | 1411 | 214 | 242 | 229 | 139 | 173 |
| | 50th | 1434 | 1434 | 1432 | 297 | 353 | 322 | 175 | 227 |
| | 75th | 1447 | 1447 | 1446 | 381 | 493 | 457 | 214 | 290 |
| | 95th | 1458 | 1458 | 1458 | 561 | 737 | 660 | 285 | 391 |
| Late | Mean | 1393 | 1396 | 1401 | 348 | 457 | 448 | 207 | 281 |
| | 5th | 1350 | 1348 | 1349 | 192 | 222 | 218 | 132 | 160 |
| | 25th | 1391 | 1390 | 1391 | 256 | 333 | 340 | 173 | 226 |
| | 50th | 1413 | 1413 | 1413 | 331 | 434 | 427 | 199 | 272 |
| | 75th | 1428 | 1428 | 1428 | 410 | 554 | 533 | 234 | 328 |
| | 95th | 1442 | 1442 | 1442 | 580 | 757 | 733 | 303 | 429 |

Table S12: Summary statistics for the distribution of epidemic length (in days) in a high transmission scenario.

| | | Response Level | | | | | | | |
|---|---|---|---|---|---|---|---|---|---|
| | | Worst-case | | | Moderate-case | | | Best-case | |
| Detection Time | Summary Stat | Ring | Geo | Best Geo | Ring | Geo | Best Geo | Ring | Geo |
| Early | Mean | 1233761 | 545611 | 527688 | 54118 | 67581 | 66513 | 21936 | 34352 |
| | 5th | 2483 | 2712 | 2385 | 1844 | 3577 | 2813 | 1970 | 3252 |
| | 25th | 1377315 | 601024 | 594814 | 11094 | 17453 | 15588 | 8090 | 12750 |
| | 50th | 1448570 | 638158 | 646300 | 35206 | 44260 | 46853 | 17069 | 26594 |
| | 75th | 1522855 | 667396 | 672711 | 85045 | 93762 | 102710 | 30635 | 48509 |
| | 95th | 1621906 | 690159 | 693893 | 171361 | 194071 | 190370 | 58485 | 86798 |
| Medium | Mean | 1334818 | 582987 | 582783 | 78756 | 100358 | 91040 | 32650 | 47747 |
| | 5th | 7478 | 6788 | 13570 | 6902 | 8367 | 6480 | 5282 | 6855 |
| | 25th | 1394835 | 612280 | 613936 | 29994 | 40256 | 37672 | 15609 | 25726 |
| | 50th | 1455580 | 647158 | 650580 | 60980 | 85947 | 77488 | 27738 | 43172 |
| | 75th | 1519072 | 666117 | 672272 | 105906 | 146627 | 129829 | 45656 | 67502 |
| | 95th | 1607922 | 686083 | 693386 | 207260 | 237159 | 221401 | 75239 | 100150 |
| Late | Mean | 1438667 | 617571 | 626262 | 131722 | 144648 | 142788 | 62335 | 75496 |
| | 5th | 1344968 | 562700 | 569100 | 32159 | 45462 | 46957 | 19796 | 31484 |
| | 25th | 1401680 | 599000 | 606000 | 78491 | 87280 | 95729 | 41525 | 53295 |
| | 50th | 1448380 | 624500 | 629000 | 120444 | 133949 | 136316 | 59758 | 73442 |
| | 75th | 1501280 | 649500 | 655000 | 170323 | 194817 | 183334 | 81297 | 94514 |
| | 95th | 1595654 | 676542 | 681375 | 271577 | 278615 | 261414 | 107970 | 124741 |

Table S13: Summary statistics for the distribution of number of vaccine doses administered in a low transmission scenario.

#### 2.2 Cost-effectiveness: deaths averted

##### 2.2.1 Moderate transmission setting

Figure S6 shows estimates of incremental net monetary benefits (INMB) calculated using deaths averted comparing targeted geographic vaccination against the current standard-of care ring vaccination within worst- and moderate-case response scenarios coupled with an improved best-case response, all within a moderate transmission setting.

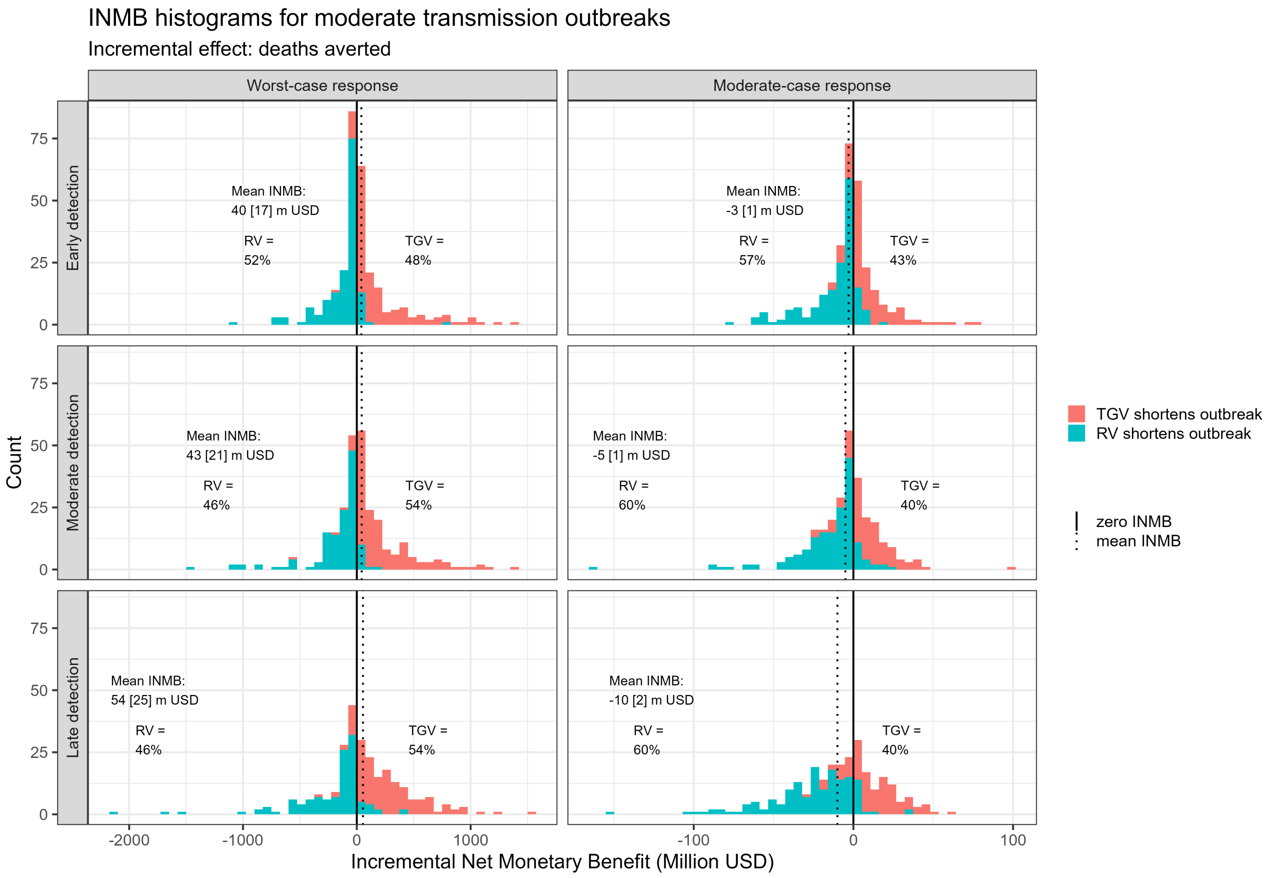


Figure S6: Incremental net monetary benefit (INMB) histograms comparing 'worst-case + best-case geo' to worst-case ring vaccination, and 'moderate-case + best-case geo' to moderate ring vaccination. Positive values of INMB (in red) indicate that TGV shortens epidemic duration more than RV, and negative values (in blue) indicate that RV shortens outbreak length more than TGV. In each subplot, the mean INMB for the given scenario is shown, along with the SEM (in brackets), and the percentage of positive and negative INMBs.

##### 2.2.2 Low transmission setting

Figure S7 shows INMBs (calculated using deaths averted) comparing geographically targeted against ring vaccination for worst-case, moderate-case, and best-case scenarios in a low transmission setting. Figure S8 shows similar figures comparing targeted geographic vaccination against ring vaccination used within worst- and moderate-case response scenarios coupled with an improved best-case response.

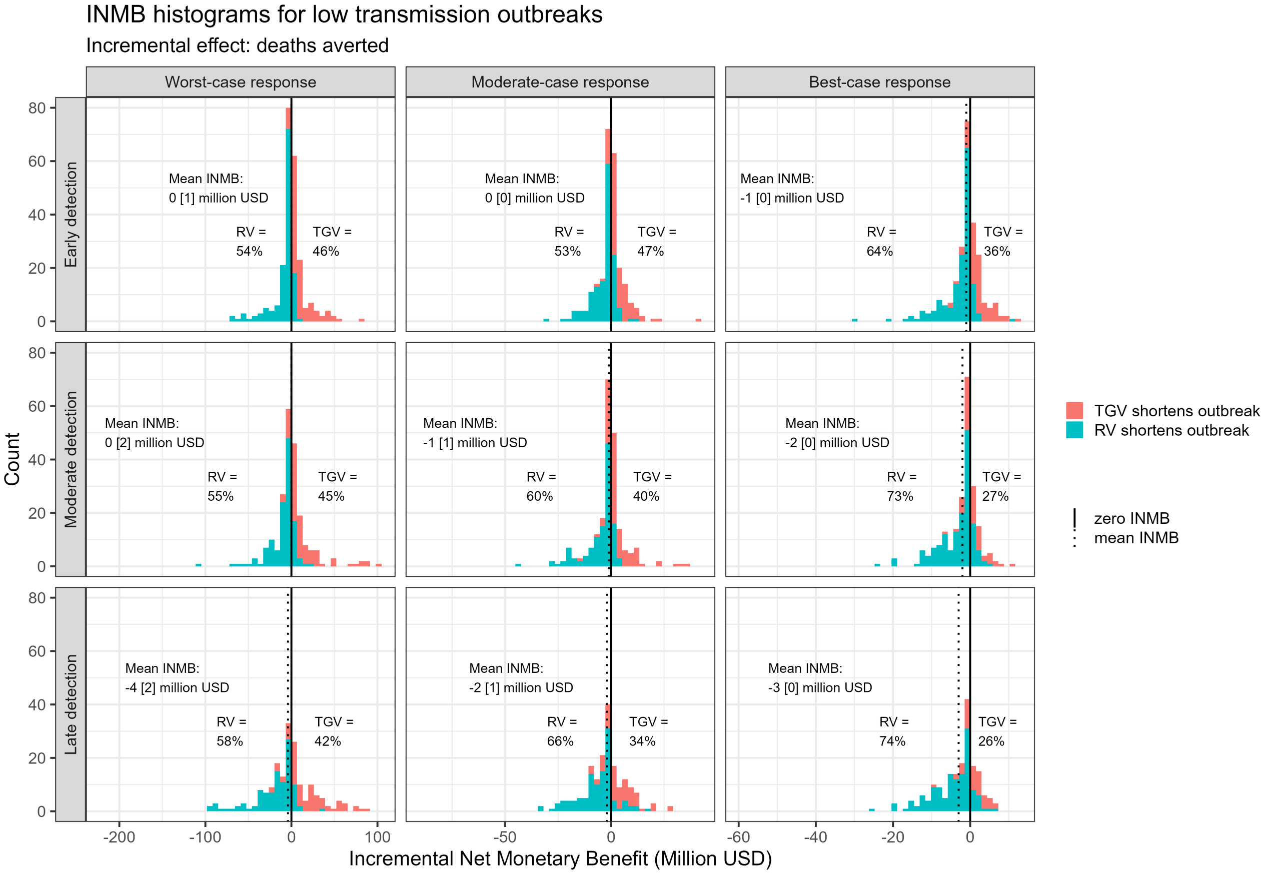


Figure S7: Histograms showing INMBs for targeted geographic vaccination relative to ring vaccination in a low transmission setting. Detection time varies across the rows (top: early, middle: medium, bottom: late) and response level increases across the columns from left to right. Positive values of INMB (in red) indicate that TGV shortens epidemic duration more than RV, and negative values (in blue) indicate that RV shortens outbreak length more than TGV. In each subplot, the mean INMB for the given scenario is shown, along with the SEM in brackets, and the percentage of positive and negative INMBs.

##### 2.2.3 High transmission setting

Figure S7 shows INMB (calculated using deaths averted) comparing geographically targeted against ring vaccination for worst-case, moderate-case, and best-case scenarios in a high transmission setting. Figure S8 and shows similar figures comparing targeted geographic vaccination against ring vaccination within worst- and moderate-case response scenarios versus coupled with an improved best-case response.

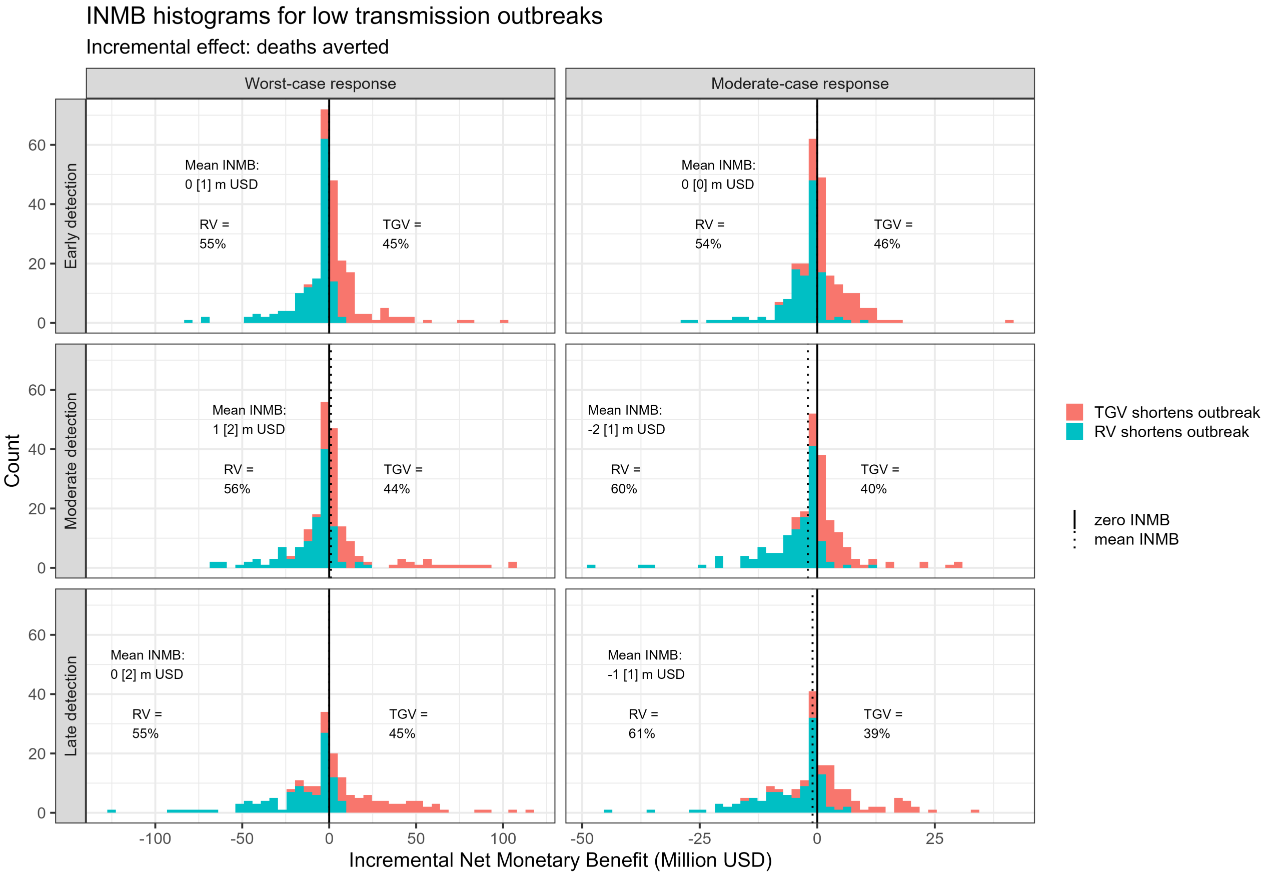


Figure S8: INMB histograms comparing 'worst-case + best geo' to worst-case ring vaccination, and 'moderate-case + best geo' to moderate-case ring vaccination in a low transmission setting. Positive values of INMB (in red) indicate that TGV shortens epidemic duration more than RV, and negative values (in blue) indicate that RV shortens outbreak length more than TGV. In each subplot, the mean INMB for the given scenario is shown, along with the SEM (in brackets), and the percentage of positive and negative INMBs.

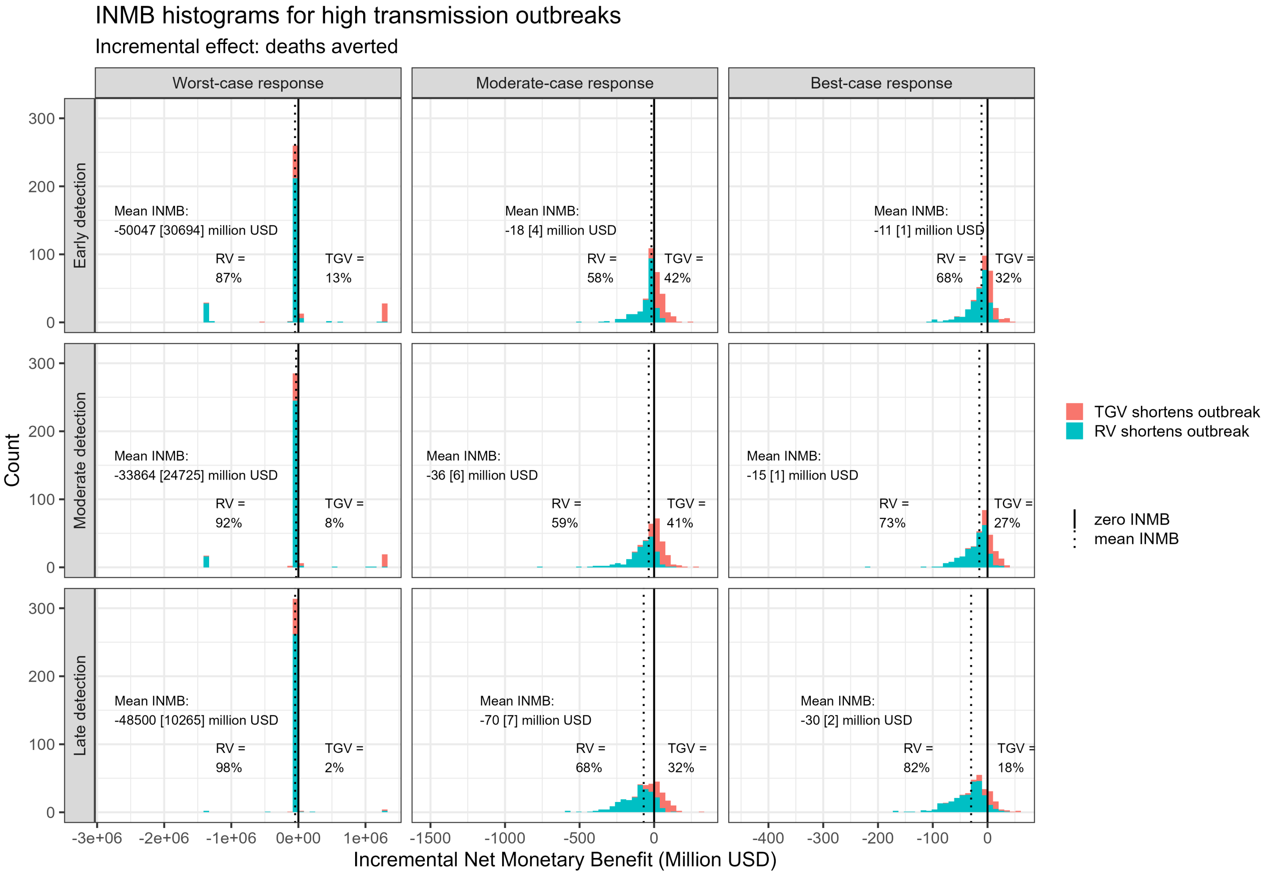


Figure S9: Histograms showing INMB for targeted geographic vaccination relative to ring vaccination in a high transmission. Detection time varies across the rows (top: early, middle: medium, bottom: late) and response level increases across the columns from left to right. Positive values of INMB (in red) indicate that TGV shortens epidemic duration more than RV, and negative values (in blue) indicate that RV shortens outbreak length more than TGV. In each subplot, the mean INMB for the given scenario is shown, along with the SEM in brackets, and the percentage of positive and negative INMBs.

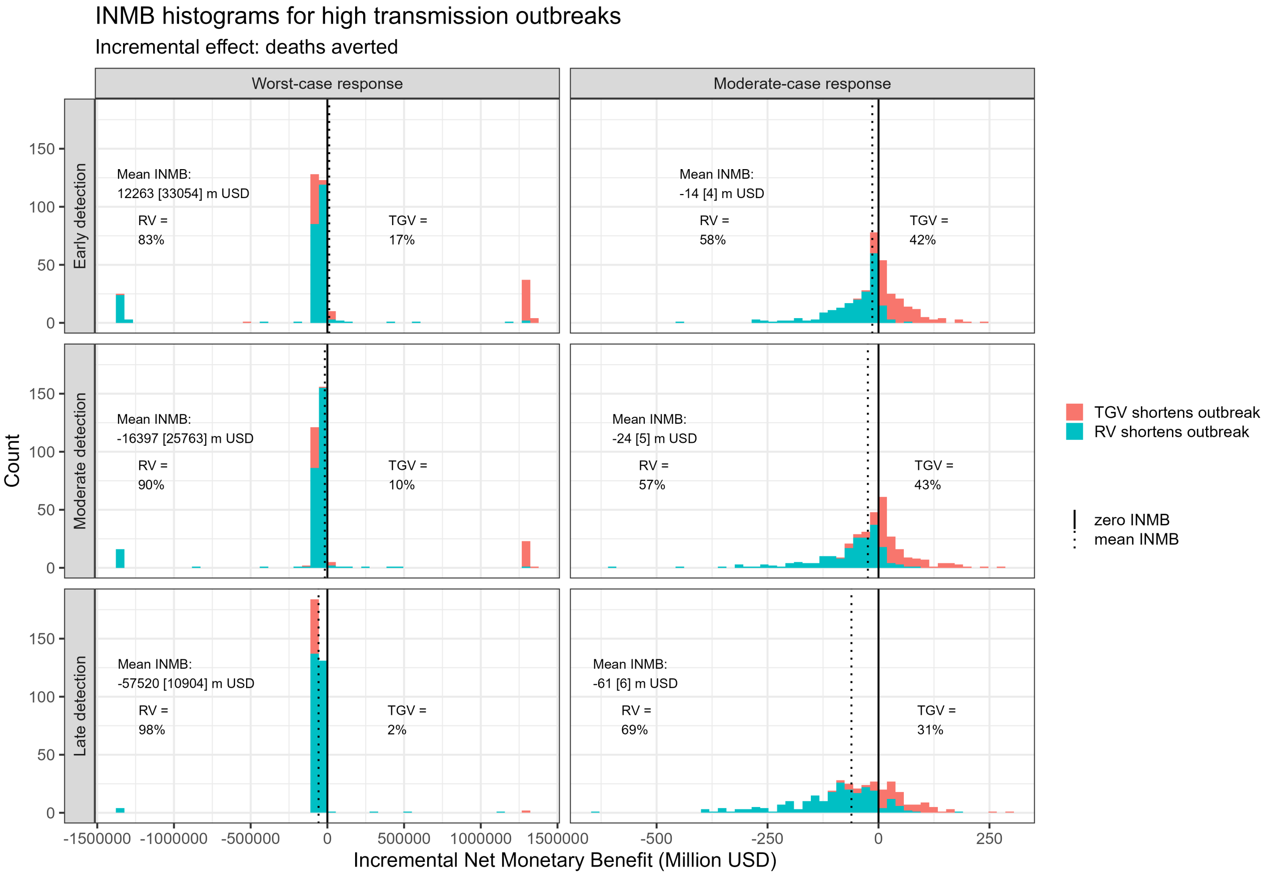


Figure S10: INMB histograms comparing 'worst-case + best geo' to worst-case ring vaccination, and 'moderate-case + best geo' to moderate-case ring vaccination in a high transmission setting. Positive values of INMB (in red) indicate that TGV shortens epidemic duration more than RV, and negative values (in blue) indicate that RV shortens outbreak length more than TGV.In each subplot, the mean INMB for the given scenario is shown, along with the SEM (in brackets), and the percentage of positive and negative INMBs.

#### 2.3 Cost-effectiveness: YLLs averted

##### 2.3.1 Moderate transmission setting

Figure S11 shows incremental net monetary benefits (INMB) calculated using YLLs averted comparing geographically targeted against ring vaccination for worst-case, moderate-case, and best-case scenarios in a moderate transmission setting. Figure S12 shows estimates of INMB (calculated using YLLs averted) comparing targeted geographic vaccination against the current standard-of care ring vaccination within worst- and moderate-case response scenarios coupled with an improved best-case response, all within a moderate transmission setting.

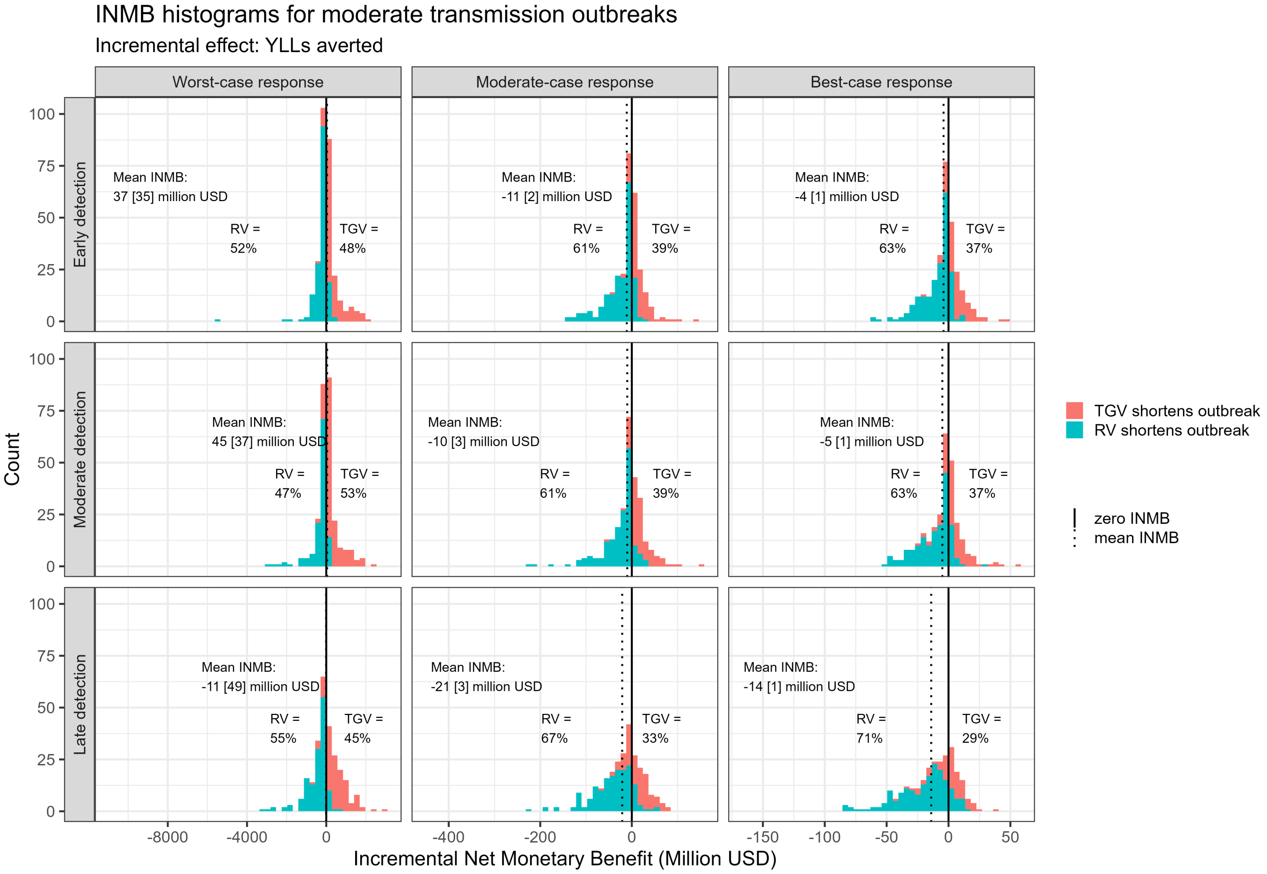


Figure S11: Histograms showing INMBs for targeted geographic vaccination relative to ring vaccination in a moderate transmission setting. Detection time varies across the rows (top: early, middle: medium, bottom: late) and response level increases across the columns from left to right. Positive values of INMB (in red) indicate that TGV shortens epidemic duration more than RV, and negative values (in blue) indicate that RV shortens outbreak length more than TGV. In each subplot, the mean INMB for the given scenario is shown, along with the SEM in brackets, and the percentage of positive and negative INMBs.

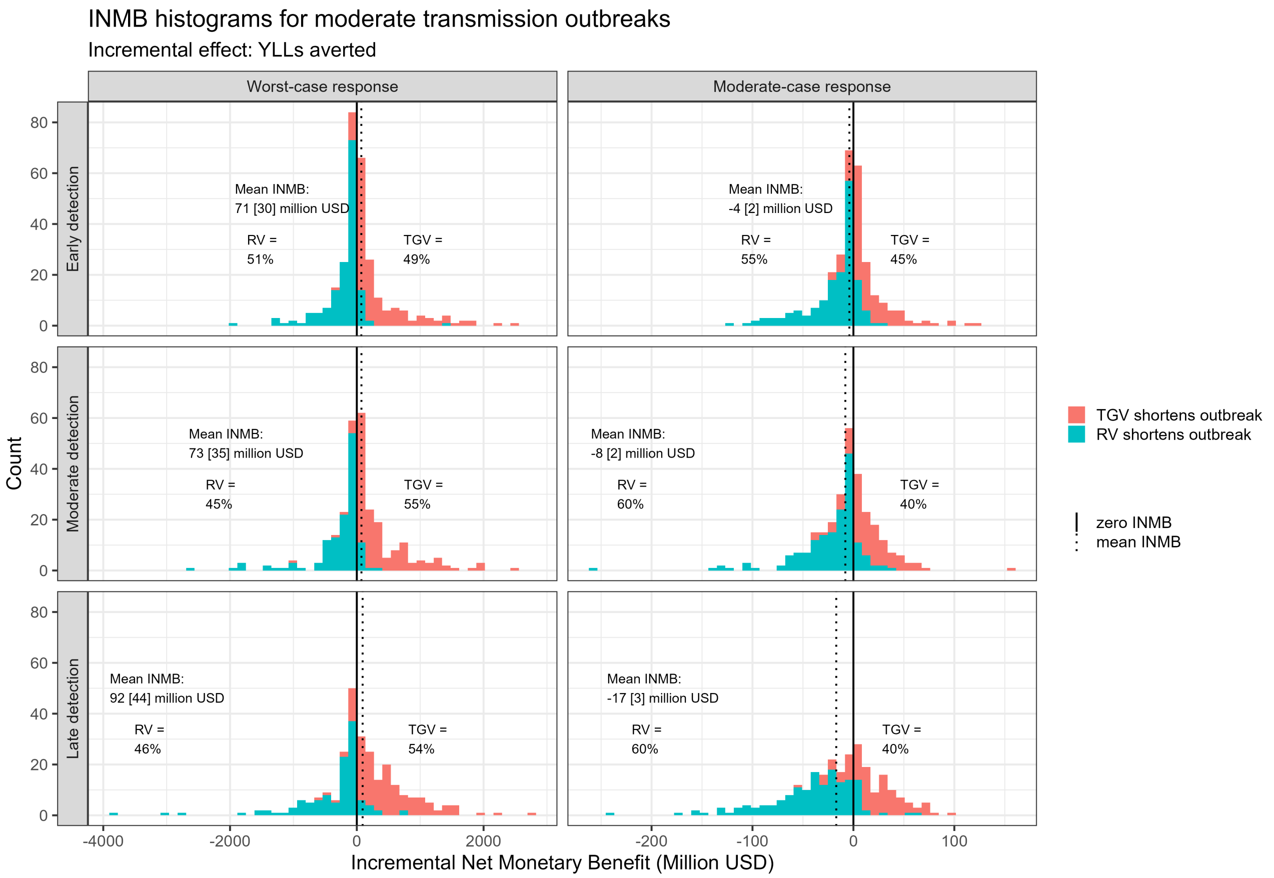


Figure S12: Incremental net monetary benefit (INMB) histograms comparing 'worst-case + best-case geo' to worst-case ring vaccination, and 'moderate-case + best-case geo' to moderate ring vaccination. Positive values of INMB (in red) indicate that TGV shortens epidemic duration more than RV, and negative values (in blue) indicate that RV shortens outbreak length more than TGV. In each subplot, the mean INMB for the given scenario is shown, along with the SEM (in brackets), and the percentage of positive and negative INMBs.

##### 2.3.2 Low transmission setting

Figure S13 shows INMBs (calculated using YLLs averted) comparing geographically targeted against ring vaccination for worst-case, moderate-case, and best-case scenarios in a low transmission setting. Figure S14 shows similar figures comparing targeted geographic vaccination against ring vaccination used within worst- and moderate-case response scenarios coupled with an improved best-case response.

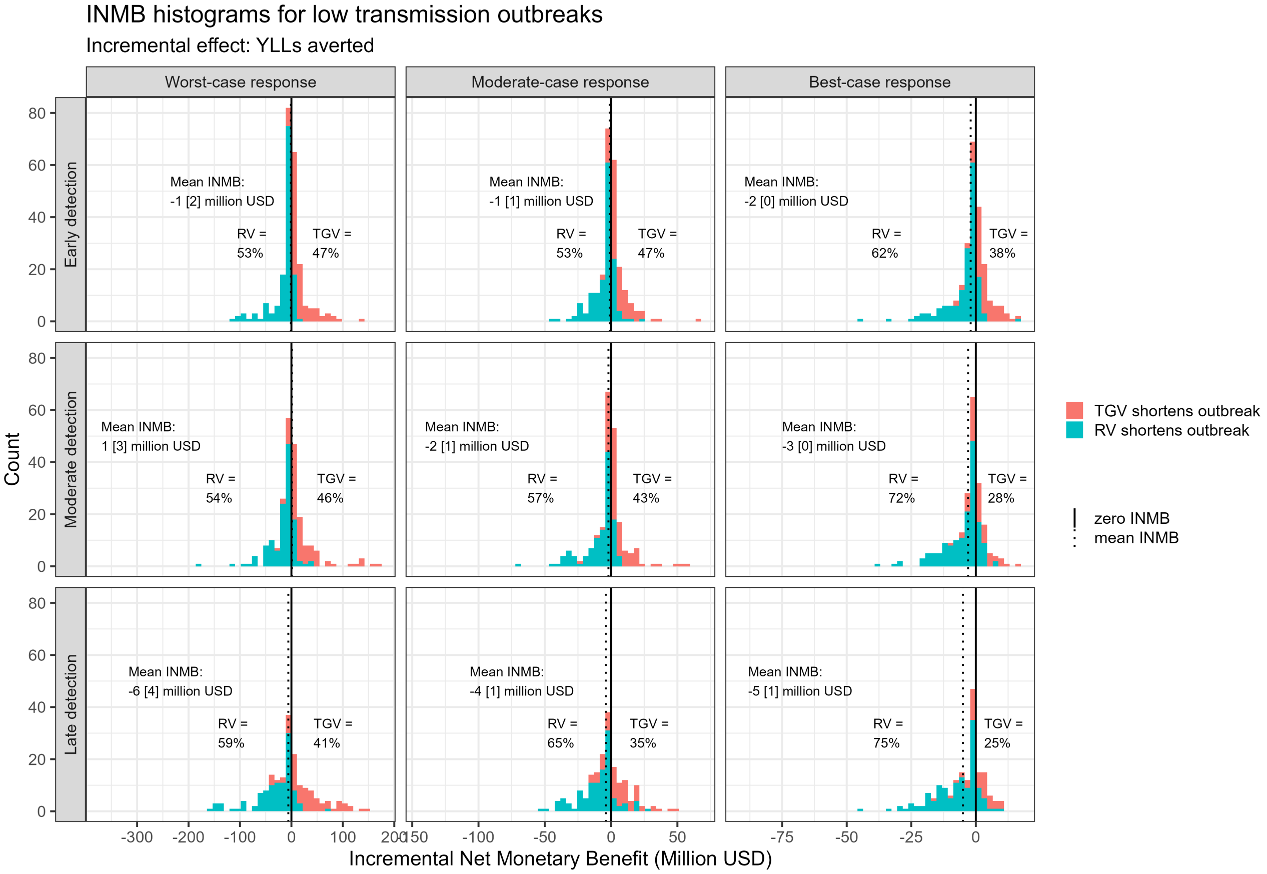


Figure S13: Histograms showing INMBs for targeted geographic vaccination relative to ring vaccination in a low transmission setting. Detection time varies across the rows (top: early, middle: medium, bottom: late) and response level increases across the columns from left to right. Positive values of INMB (in red) indicate that TGV shortens epidemic duration more than RV, and negative values (in blue) indicate that RV shortens outbreak length more than TGV. In each subplot, the mean INMB for the given scenario is shown, along with the SEM in brackets, and the percentage of positive and negative INMBs.

##### 2.3.3 High transmission setting

Figure S15 shows INMB (calculated using YLLs averted) comparing geographically targeted against ring vaccination for worst-case, moderate-case, and best-case scenarios in a high transmission setting. Figure S16 and shows similar figures comparing targeted geographic vaccination against ring vaccination within worst- and moderate-case response scenarios versus coupled with an improved best-case response.

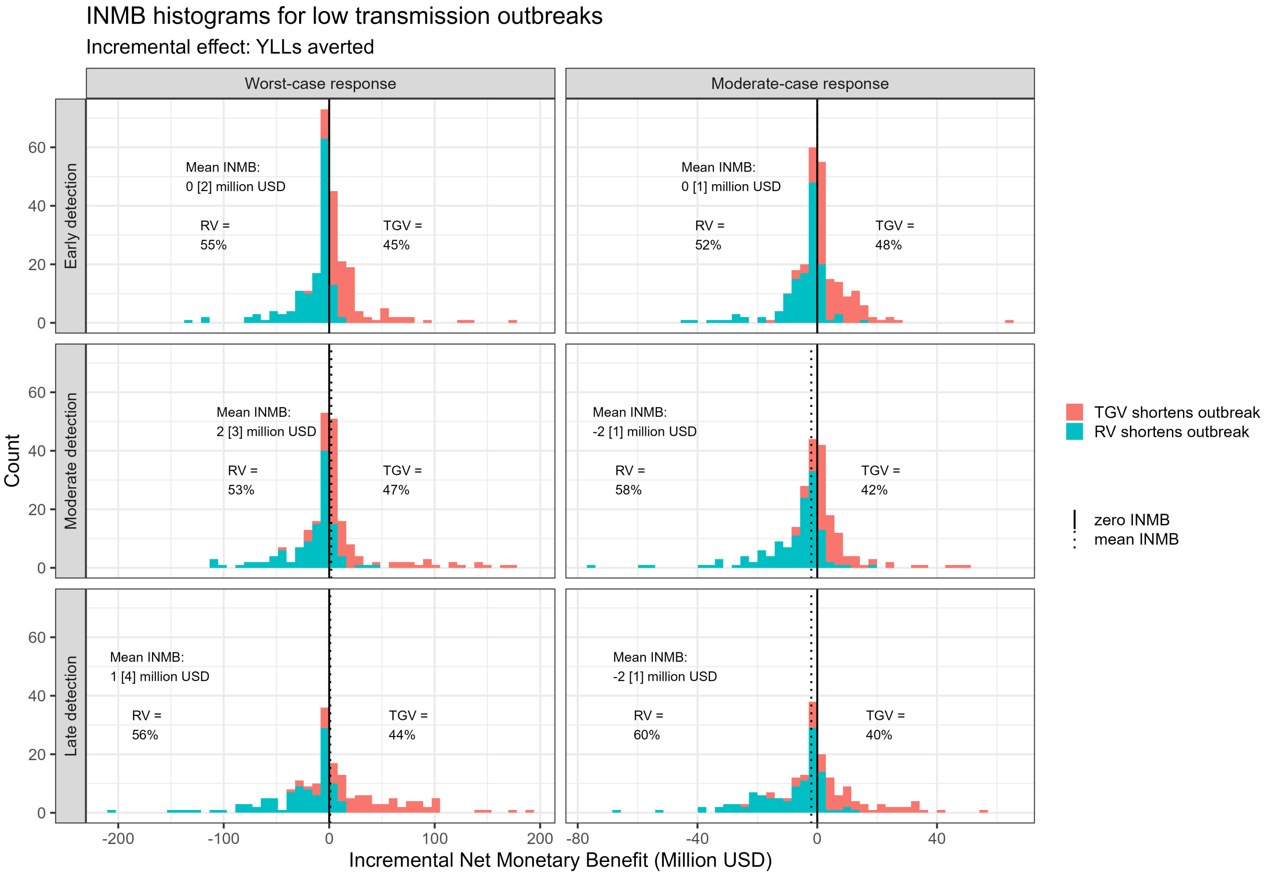


Figure S14: INMB histograms comparing 'worst-case + best geo' to worst-case ring vaccination, and 'moderate-case + best geo' to moderate-case ring vaccination in a low transmission setting. Positive values of INMB (in red) indicate that TGV shortens epidemic duration more than RV, and negative values (in blue) indicate that RV shortens outbreak length more than TGV. In each subplot, the mean INMB for the given scenario is shown, along with the SEM (in brackets), and the percentage of positive and negative INMBs.

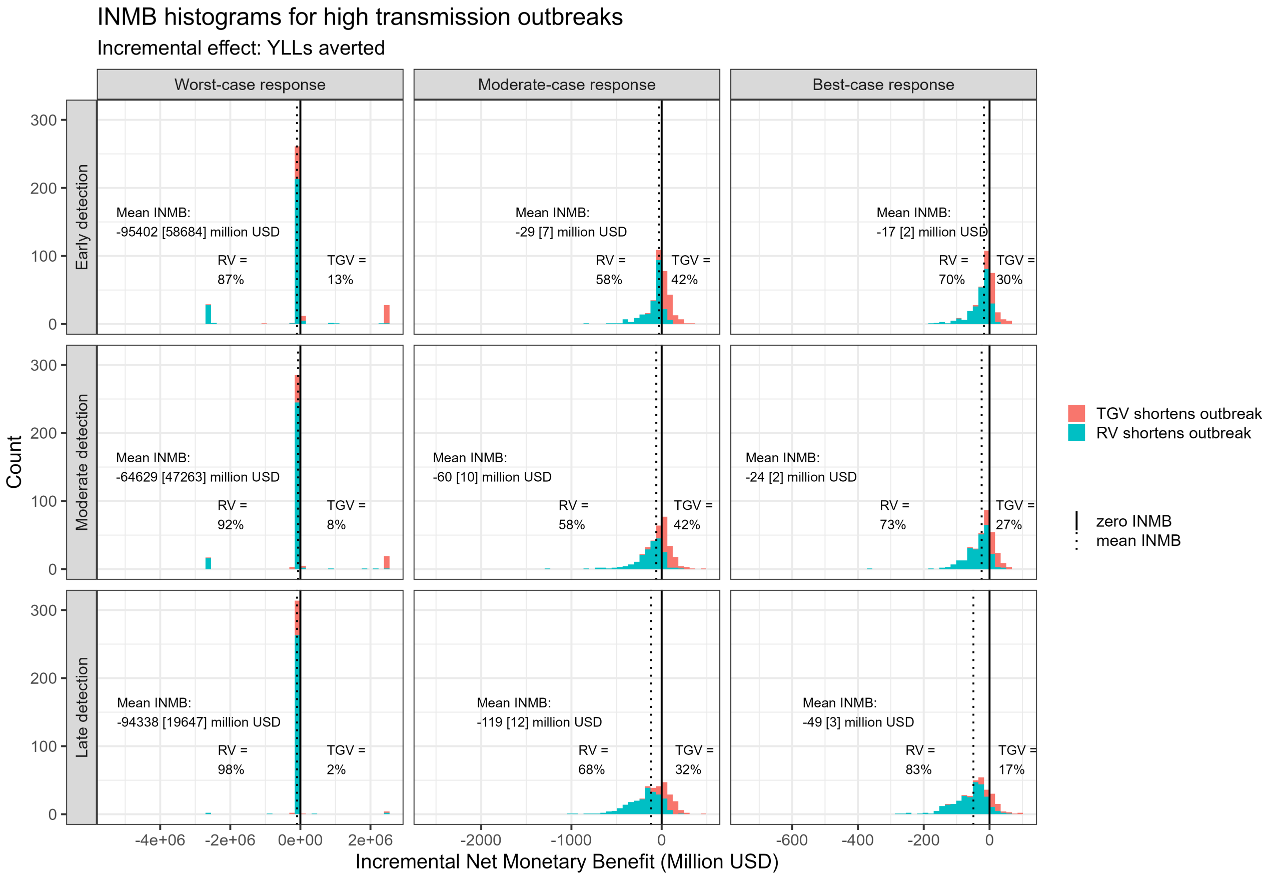


Figure S15: Histograms showing INMB for targeted geographic vaccination relative to ring vaccination in a high transmission. Detection time varies across the rows (top: early, middle: medium, bottom: late) and response level increases across the columns from left to right. Positive values of INMB (in red) indicate that TGV shortens epidemic duration more than RV, and negative values (in blue) indicate that RV shortens outbreak length more than TGV. In each subplot, the mean INMB for the given scenario is shown, along with the SEM in brackets, and the percentage of positive and negative INMBs.

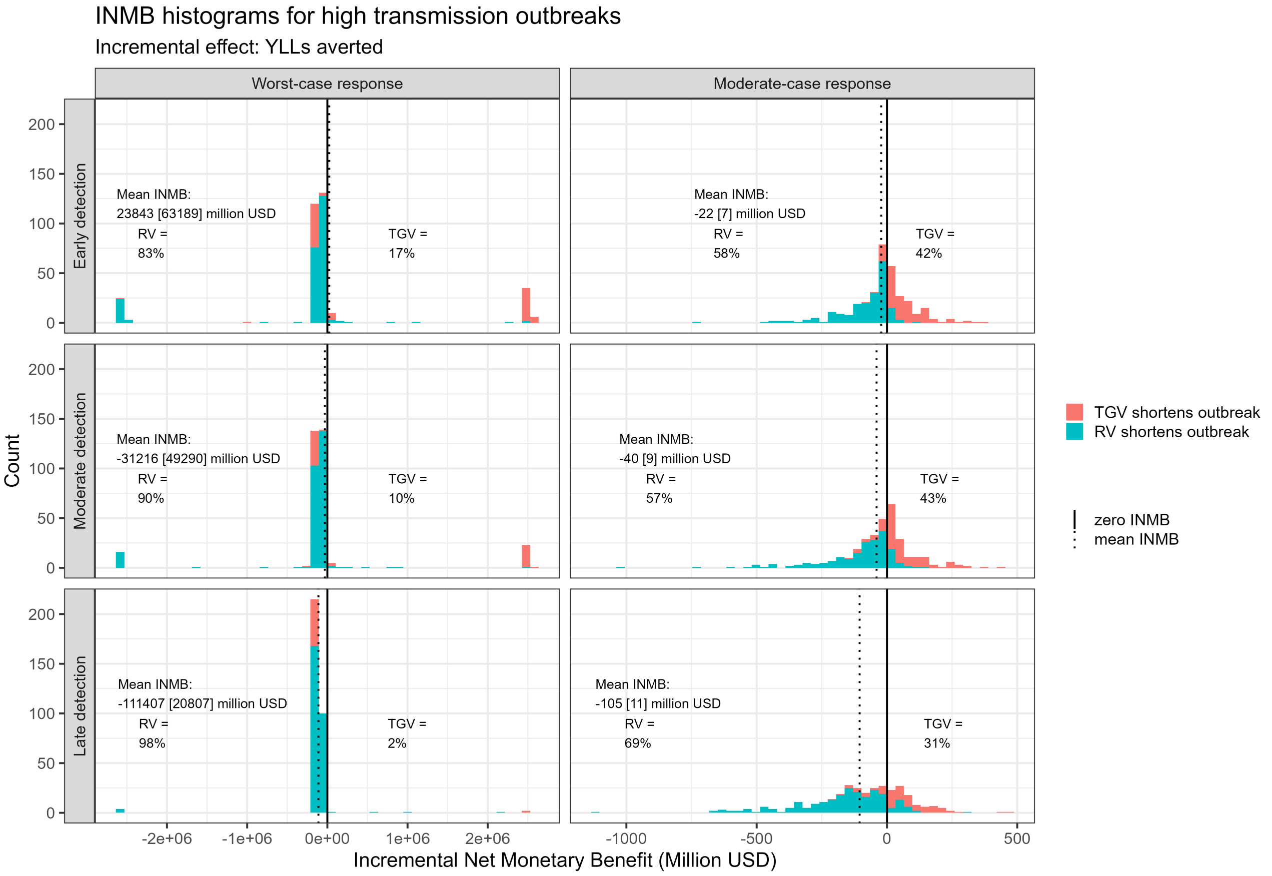


Figure S16: INMB histograms comparing 'worst-case + best geo' to worst-case ring vaccination, and 'moderate-case + best geo' to moderate-case ring vaccination in a high transmission setting. Positive values of INMB (in red) indicate that TGV shortens epidemic duration more than RV, and negative values (in blue) indicate that RV shortens outbreak length more than TGV.In each subplot, the mean INMB for the given scenario is shown, along with the SEM (in brackets), and the percentage of positive and negative INMBs.

#### 2.4 Cost-effectiveness: discounted YLLs averted

##### 2.4.1 Moderate transmission setting

Figure S17 shows incremental net monetary benefits (INMB) calculated using discounted YLLs (DYLLs) averted comparing geographically targeted against ring vaccination for worst-case, moderate-case, and best-case scenarios in a moderate transmission setting. Figure S18 shows estimates of INMB (calculated using DYLLs averted) comparing targeted geographic vaccination against the current standard-of care ring vaccination within worst- and moderate-case response scenarios coupled with an improved best-case response, all within a moderate transmission setting.

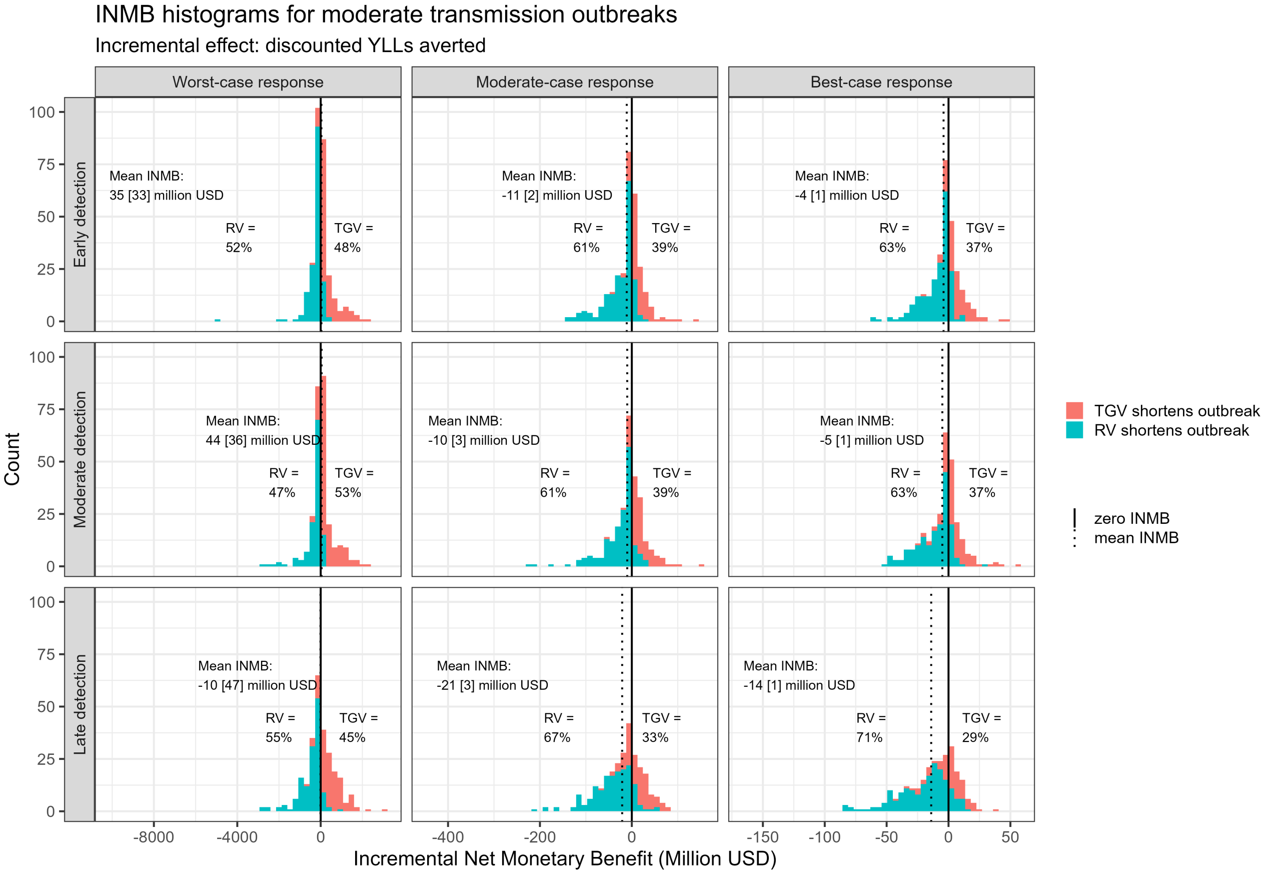


Figure S17: Histograms showing INMBs for targeted geographic vaccination relative to ring vaccination in a moderate transmission setting. Detection time varies across the rows (top: early, middle: medium, bottom: late) and response level increases across the columns from left to right. Positive values of INMB (in red) indicate that TGV shortens epidemic duration more than RV, and negative values (in blue) indicate that RV shortens outbreak length more than TGV. In each subplot, the mean INMB for the given scenario is shown, along with the SEM in brackets, and the percentage of positive and negative INMBs.

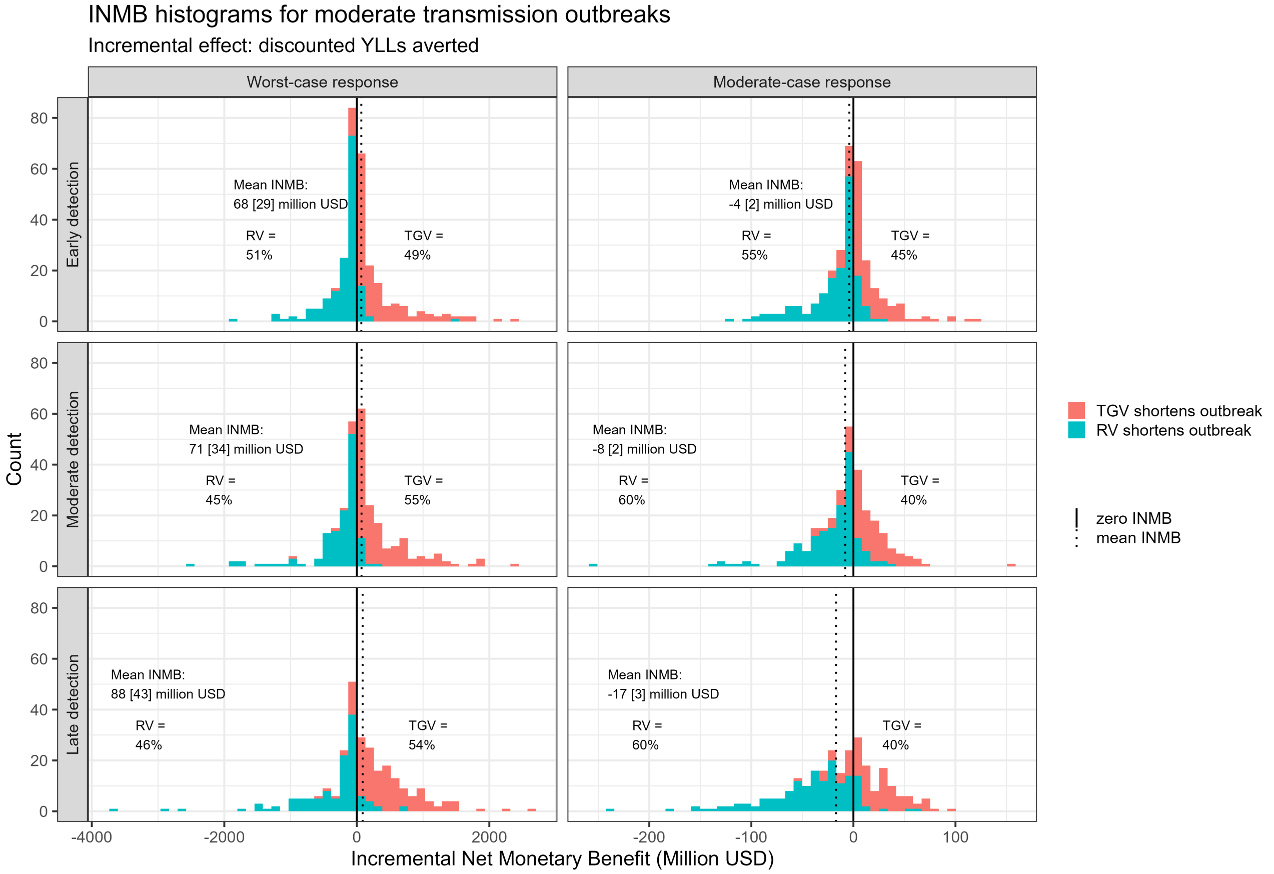


Figure S18: Incremental net monetary benefit (INMB) histograms comparing 'worst-case + best-case geo' to worst-case ring vaccination, and 'moderate-case + best-case geo' to moderate ring vaccination. Positive values of INMB (in red) indicate that TGV shortens epidemic duration more than RV, and negative values (in blue) indicate that RV shortens outbreak length more than TGV. In each subplot, the mean INMB for the given scenario is shown, along with the SEM (in brackets), and the percentage of positive and negative INMBs.

##### 2.4.2 Low transmission setting

Figure S19 shows INMBs (calculated using DYLLs averted) comparing geographically targeted against ring vaccination for worst-case, moderate-case, and best-case scenarios in a low transmission setting. Figure S20 shows similar figures comparing targeted geographic vaccination against ring vaccination used within worst- and moderate-case response scenarios coupled with an improved best-case response.

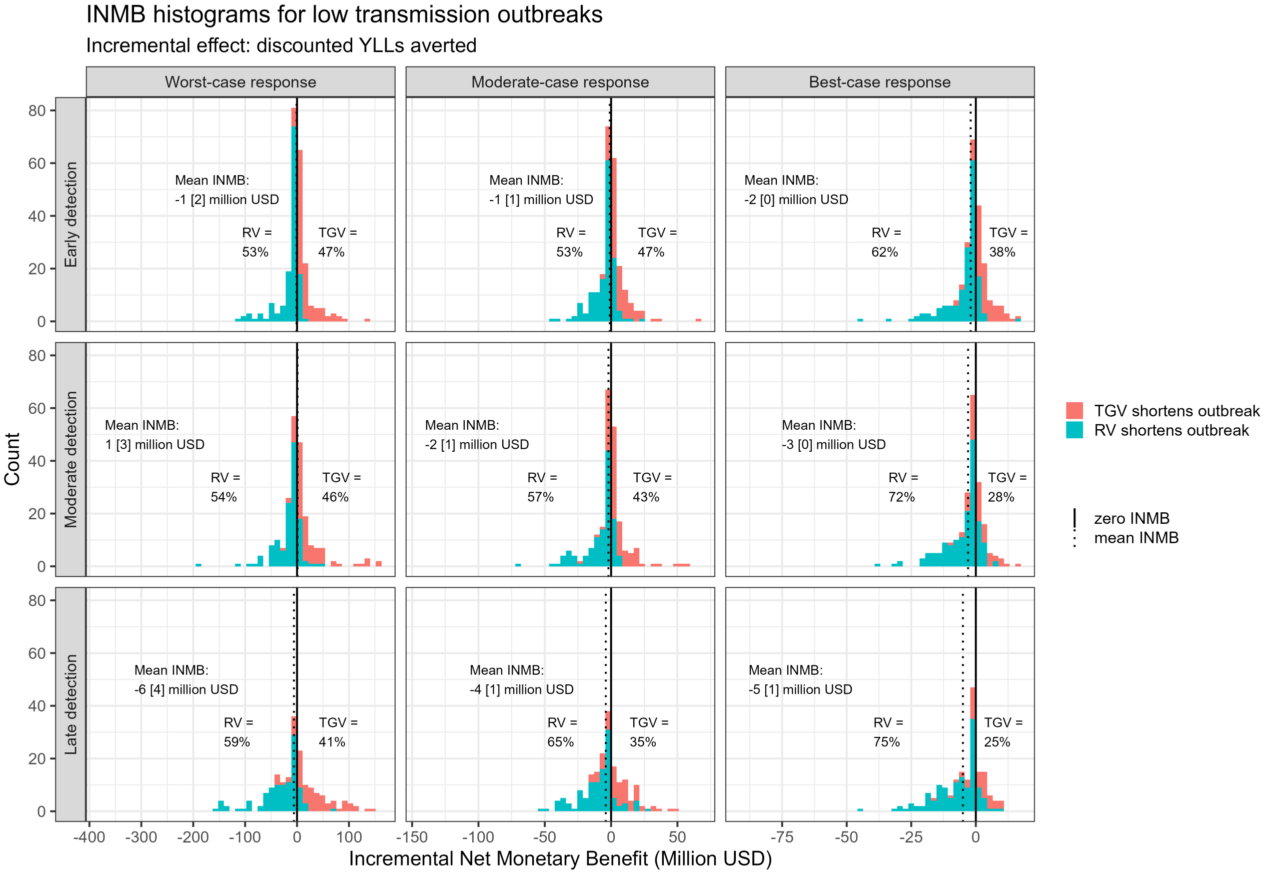


Figure S19: Histograms showing INMBs for targeted geographic vaccination relative to ring vaccination in a low transmission setting. Detection time varies across the rows (top: early, middle: medium, bottom: late) and response level increases across the columns from left to right. Positive values of INMB (in red) indicate that TGV shortens epidemic duration more than RV, and negative values (in blue) indicate that RV shortens outbreak length more than TGV. In each subplot, the mean INMB for the given scenario is shown, along with the SEM in brackets, and the percentage of positive and negative INMBs.

##### 2.4.3 High transmission setting

Figure S21 shows INMB (calculated using DYLLs averted) comparing geographically targeted against ring vaccination for worst-case, moderate-case, and best-case scenarios in a high transmission setting. Figure S22 and shows similar figures comparing targeted geographic vaccination against ring vaccination within worst- and moderate-case response scenarios versus coupled with an improved best-case response.

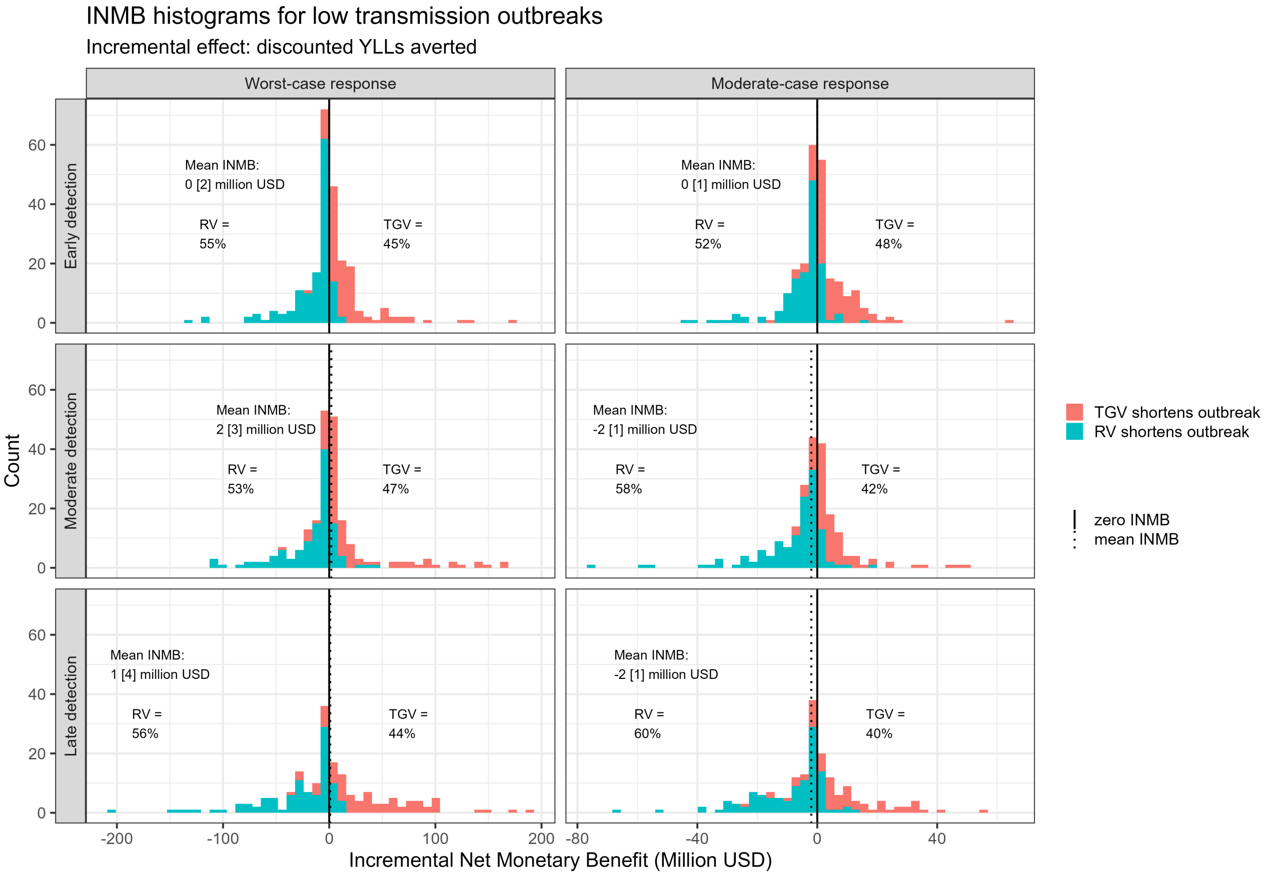


Figure S20: INMB histograms comparing 'worst-case + best geo' to worst-case ring vaccination, and 'moderate-case + best geo' to moderate-case ring vaccination in a low transmission setting. Positive values of INMB (in red) indicate that TGV shortens epidemic duration more than RV, and negative values (in blue) indicate that RV shortens outbreak length more than TGV. In each subplot, the mean INMB for the given scenario is shown, along with the SEM (in brackets), and the percentage of positive and negative INMBs.

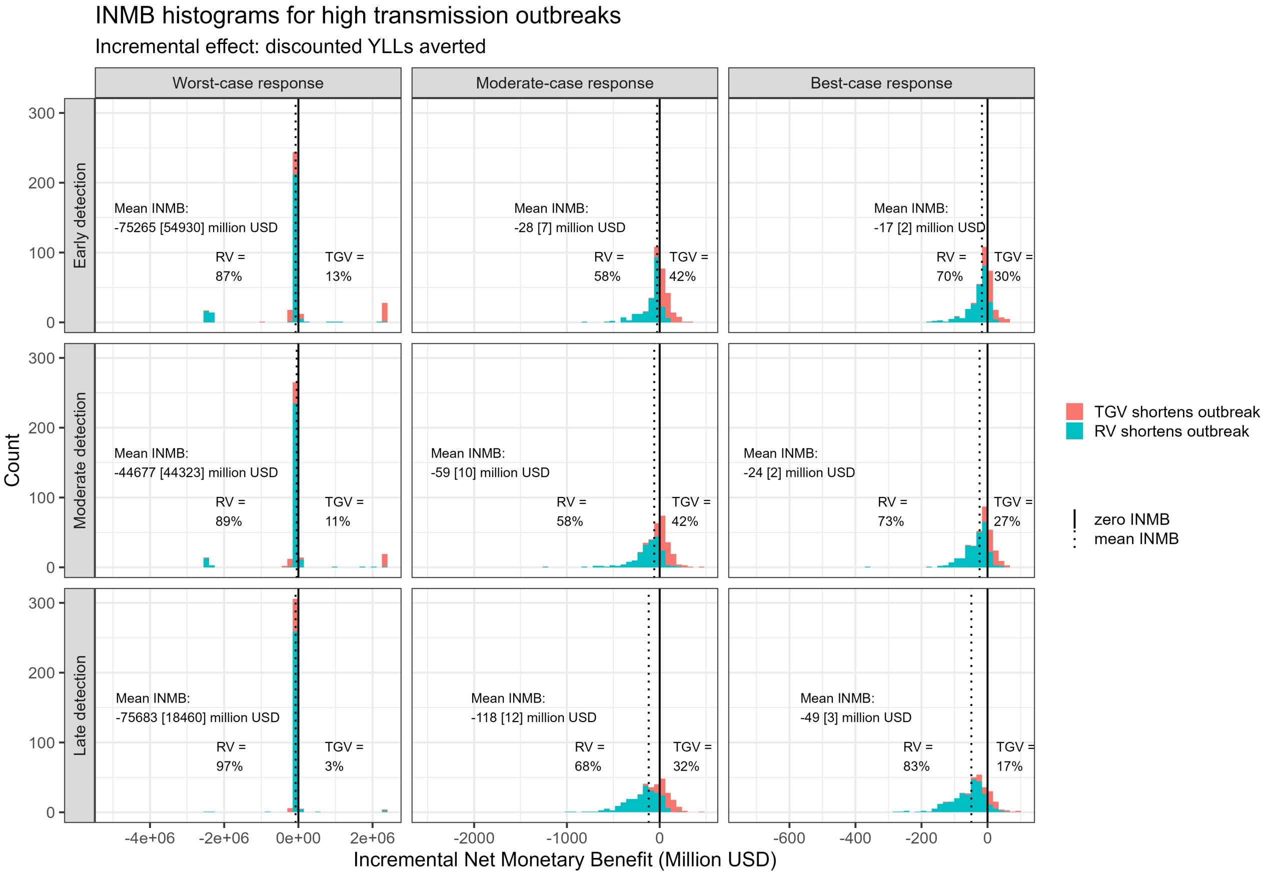


Figure S21: Histograms showing INMB for targeted geographic vaccination relative to ring vaccination in a high transmission. Detection time varies across the rows (top: early, middle: medium, bottom: late) and response level increases across the columns from left to right. Positive values of INMB (in red) indicate that TGV shortens epidemic duration more than RV, and negative values (in blue) indicate that RV shortens outbreak length more than TGV. In each subplot, the mean INMB for the given scenario is shown, along with the SEM in brackets, and the percentage of positive and negative INMBs.

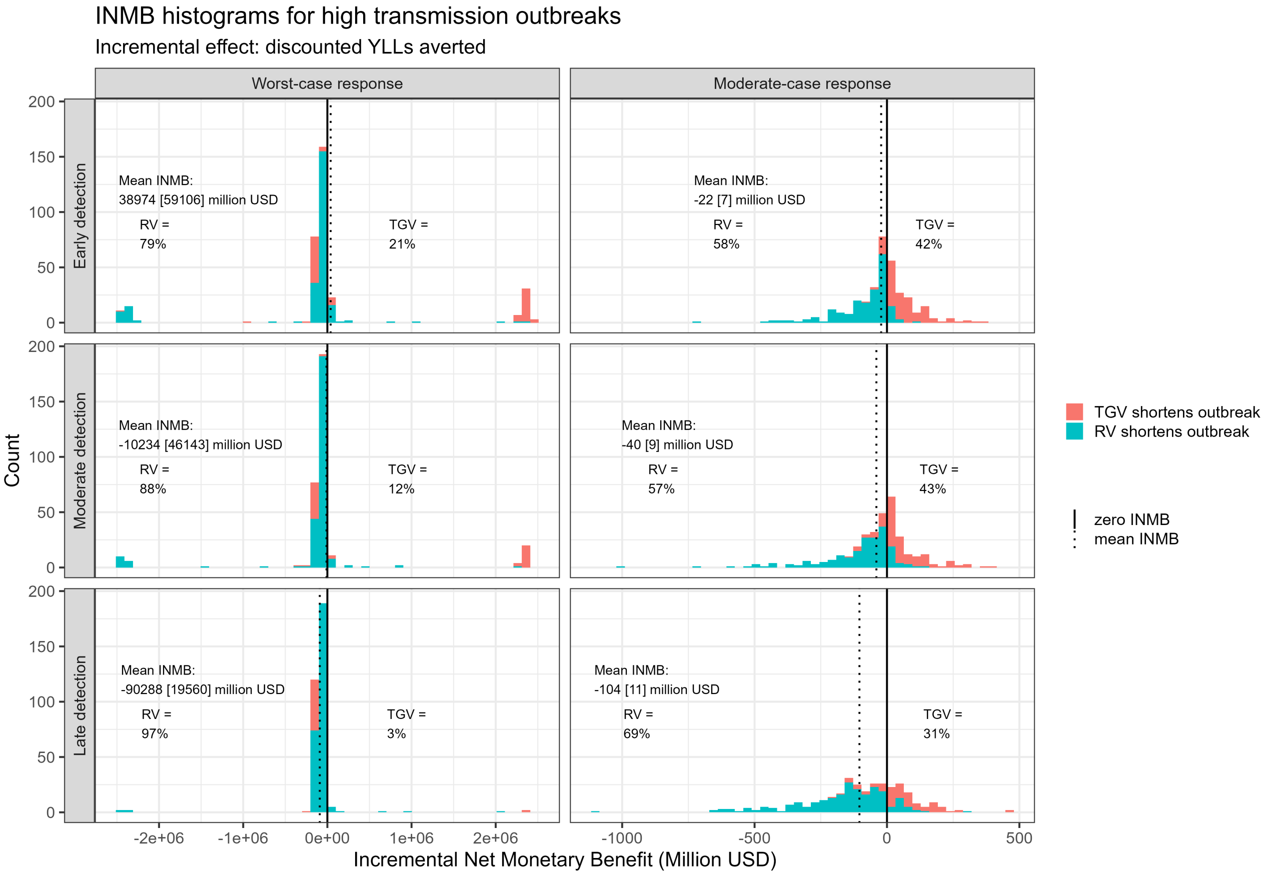


Figure S22: INMB histograms comparing 'worst-case + best geo' to worst-case ring vaccination, and 'moderate-case + best geo' to moderate-case ring vaccination in a high transmission setting. Positive values of INMB (in red) indicate that TGV shortens epidemic duration more than RV, and negative values (in blue) indicate that RV shortens outbreak length more than TGV.In each subplot, the mean INMB for the given scenario is shown, along with the SEM (in brackets), and the percentage of positive and negative INMBs.
